# *ELF5* is a respiratory epithelial cell-specific risk gene for severe COVID-19

**DOI:** 10.1101/2022.01.17.22269283

**Authors:** Maik Pietzner, Robert Lorenz Chua, Eleanor Wheeler, Katharina Jechow, Helena Radbruch, Saskia Trump, Bettina Heidecker, Frank L. Heppner, Roland Eils, Marcus A. Mall, Leif-Erik Sander, Irina Lehmann, Sören Lukassen, Nick Wareham, Christian Conrad, Claudia Langenberg

**Affiliations:** Computational Medicine, Berlin Institute of Health (BIH) at Charité – Universitätsmedizin Berlin, Germany; MRC Epidemiology Unit, University of Cambridge, Cambridge, UK; Center for Digital Health, Berlin Institute of Health (BIH) at Charité – Universitätsmedizin Berlin, Germany; Department of Neuropathology, Charité – Universitätsmedizin Berlin, corporate member of Freie Universität Berlin und Humboldt-Universität zu Berlin, Berlin, Germany; Molecular Epidemiology Unit, Charité - Universitätsmedizin Berlin, corporate member of Freie Universität Berlin, Humboldt-Universität zu Berlin and Berlin Institute of Health (BIH), Berlin, Germany; Department of Cardiology, Charité – Universitätsmedizin Berlin, corporate member of Freie Universität Berlin und Humboldt-Universität zu Berlin, Berlin, Germany; Cluster of Excellence, NeuroCure, Berlin, Germany; German Center for Neurodegenerative Diseases (DZNE) Berlin, Berlin, Germany; Health Data Science Unit, Heidelberg University Hospital and BioQuant, Heidelberg, Germany; German Center for Lung Research (DZL), associated partner site, Augustenburger Platz 1, 13353 Berlin, Germany; Department of Pediatric Respiratory Medicine, Immunology and Critical Care Medicine, Charité-Universitätsmedizin Berlin, corporate member of Freie Universität Berlin and Humboldt-Universität zu Berlin, Berlin, Germany; Berlin Institute of Health at Charité – Universitätsmedizin Berlin, Berlin, Germany; Department of Infectious Diseases and Respiratory Medicine, Charité - Universitätsmedizin Berlin, corporate member of Freie Universität Berlin, Humboldt-Universität zu Berlin, and Berlin Institute of Health (BIH), Berlin, Germany

## Abstract

Despite two years of intense global research activity, host genetic factors that predispose to a poorer prognosis and severe course of COVID-19 infection remain poorly understood. Here, we identified eight candidate protein mediators of COVID-19 outcomes by establishing a shared genetic architecture at protein-coding loci using large-scale human genetic studies. The transcription factor ELF5 (*ELF5*) showed robust and directionally consistent associations across different outcome definitions, including a >4-fold higher risk (odds ratio: 4.85; 95%-CI: 2.65-8.89; p-value<3.1×10^−7^) for severe COVID-19 per 1 s.d. higher genetically predicted plasma ELF5. We show that *ELF5* is specifically expressed in epithelial cells of the respiratory system, such as secretory and alveolar type 2 cells, using single-cell RNA sequencing and immunohistochemistry. These cells are also likely targets of SARS-CoV-2 by colocalisation with key host factors, including *ACE2* and *TMPRSS2*. We also observed a 25% reduced risk of severe COVID-19 per 1 s.d. higher genetically predicted plasma G-CSF, a finding corroborated by a clinical trial of recombinant human G-CSF in COVID-19 patients with lymphopenia reporting a lower number of patients developing critical illness and death. In summary, large-scale human genetic studies together with gene expression at single-cell resolution highlight *ELF5* as a novel risk gene for COVID-19 prognosis, supporting a role of epithelial cells of the respiratory system in the adverse host response to SARS-CoV-2.

## INTRODUCTION

The COVID-19 pandemic, caused by the coronavirus SARS-CoV-2, has overwhelmed health care systems all over the world and caused more than 5.1 million deaths. The unprecedented pace of vaccine development, approval, and administration^1^, has strongly reduced hospitalisations and prevented hundreds of thousands of deaths^2^, and only achieving population-wide immunity will end the pandemic. However, it is unclear how long immunisation from vaccines or natural infection will last^3^ and hospitalisation and death tolls remain high due to various factors, including the evolution of novel SARS-CoV-2 variants^4–6^ and the missing availability of vaccines in low- and middle-income countries^7^, requiring persistent efforts to identify host factors that predispose to poor outcomes.

So far, older age, male sex, smoking, obesity, social deprivation, ethnicity, and a high burden of pre-existing conditions have been consistently identified as risk factors for a poor prognosis among COVID-19 patients^8–11^. However, a severe disease course, including hospitalisations and fatal outcomes also occur in otherwise low-risk patients. Further, the biology underlying disease progression and fatal outcomes remains largely unknown, with observational studies unable to dissect cause from consequence. Common variation in the human genome has now been robustly linked to a higher susceptibility to severe COVID-19 outcomes^12–14^, offering novel and orthogonal insights to deep molecular profiling studies in patients employing single cell sequencing or immunoprofiling^15–20^. For example, common variants at 3p21.31 (possibly mapping to *LZTFL1* or *SLC6A20*) or 12q24.13 (likely *OAS1*) confer 30%-110% higher risk for severe outcomes of COVID-19^12–14^, with suggested roles in alveolar type 2 (AT2) cells^16,21^ or modulation of the host immune response^22,23^. However, a major obstacle to clinical translation of these findings is the identification of the causal genes through which risk loci mediate their effect. Further, incorporating gene or protein expression quantitative trait loci (QTLs) via statistical colocalisation or Mendelian randomization can highlight additional candidate genes^22,24–28^. Both techniques make use of a causal chain of events. Firstly, alleles are allocated at random at conception providing the opportunity to use them as instruments for causal inference. Secondly, genetic variants near protein-encoding loci (cis-pQTLs) that associate with protein levels in healthy individuals before viral exposure can serve as instruments for lifelong exposure to higher or lower protein levels. Therefore, establishing that the same genetic signal associates with protein levels and a poor prognosis of COVID-19 provides strong evidence for a causal role of the protein in the aetiology of the disease. This is particularly relevant for COVID-19, which is characterized by a hyperimmune response and a profound impact on the plasma proteome^29^ limiting insights from ad hoc cross-sectional proteomic studies. Such genetically informed strategies have already identified potential druggable targets, including *ACE2*^24^, or modulators of the immune response such as OAS1^22^.

Here, we present a proteome-wide colocalisation screen, incorporating cis-pQTLs for more than 2,300 protein targets across two different platforms, to identify proteomic modulators of COVID-19 prognosis using genome-wide summary statistics for four different outcome definitions (ranging from susceptibility to severity) as released by the COVID-19 Host Genetics Initiative^30^ (https://www.covid19hg.org/, release 6) (**Fig. 1**). We demonstrate the ability of genetically informed plasma proteomics to identify causal genes and proteins for severe COVID-19 and pinpoint the responsible cell types through integration of orthogonal data.

**Figure 1.**
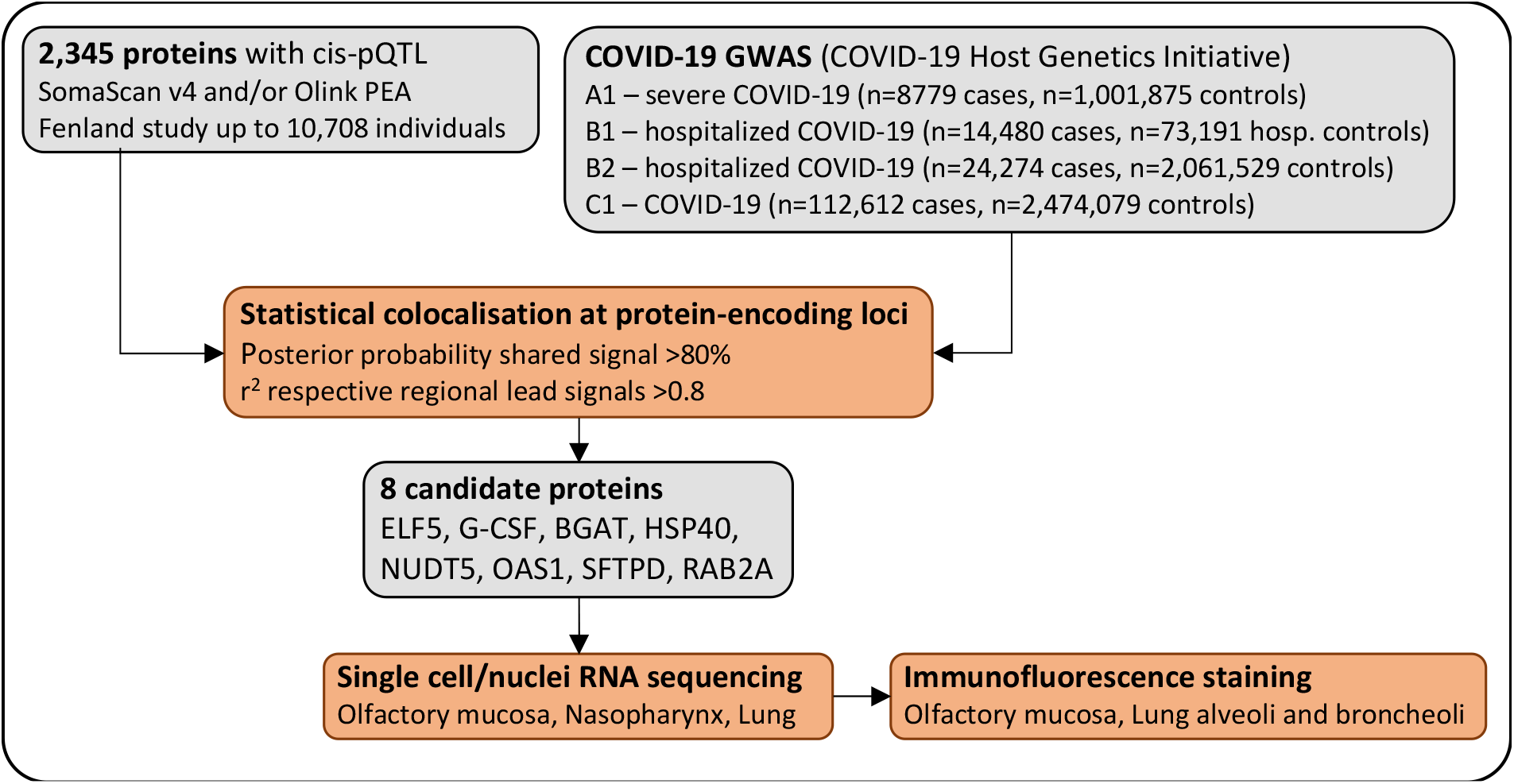
Flowchart of the study design. We tested whether the same genetic signal that associated with higher/lower protein abundances also associated with higher/lower risk for COVID-19 based on cis protein quantitative trait loci (cis-pQTLs). We further investigated expression of targeted proteins/genes using single cell and single nuclei RNA sequencing in samples of the respiratory system and confirmed expression of the most robust candidate ELF5 using immunofluorescence staining.

## RESULTS

### Putative protein mediators of disease risk

We identified 6 previously unknown candidate causal proteins, in addition to replicating the established candidates BGAT (encoded by *ABO*) and OAS1 (encoded by *OAS1*)^22,24,25^, by systematically testing for a shared genetic architecture at protein coding loci (±500kb) for 2375 protein targets (see **Methods**) and COVID-19^14^ outcomes using statistical colocalisation (**Supplementary Table 1, Methods**).

Of the 6 novel candidates, ELF5 showed significant and directionally consistent associations across all four COVID-19 outcomes included as part of the COVID-19 Host Genetics Initiative (https://www.covid19hg.org/)^14^, with strongest effects for more severe outcomes (**Fig. 2 and Supplementary Table 2**) and strong evidence for a shared genetic signal for outcomes indicating a poorer prognosis (hospitalisation and severe COVID-19; Posterior probability (PP)>80%). For example, a 1 s.d. increase in genetically predicted ELF5 plasma abundances was associated with an almost 5-fold higher risk for severe COVID-19 (odds ratio: 4.85; 95%-CI: 2.65-8.89; p-value<3.1×10^−7^) in a single-instrument Mendelian randomization (MR) analysis using the lead cis protein quantitative trait locus (cis-pQTL) as the genetic instrument (**Fig. 2**, see **Methods**).

**Figure 2.**
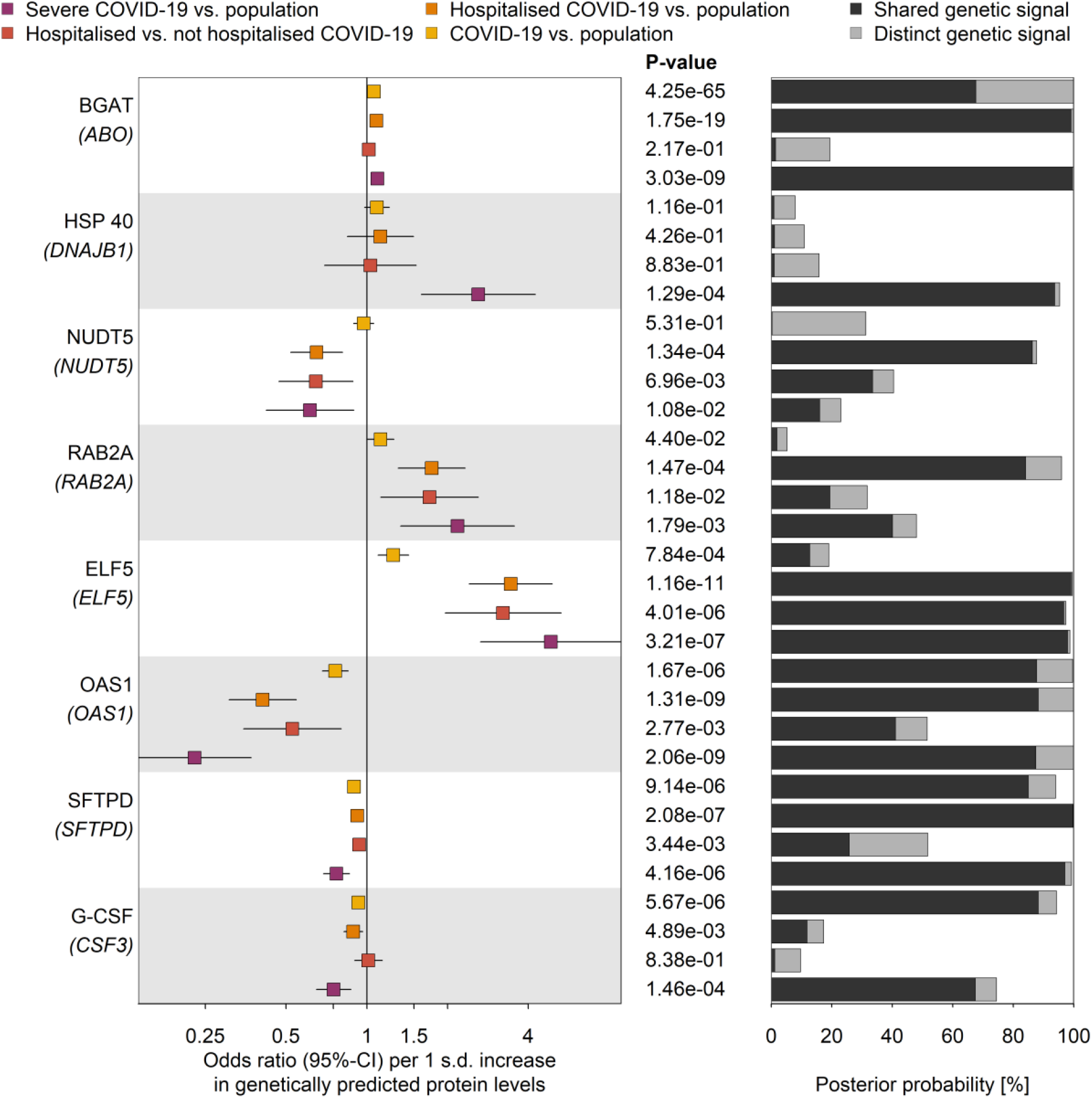
Proteins genetically linked to various COVID-19 outcomes. Odds ratios and 95%-CIs for the genetically predicted effect of protein levels on four different outcome definitions and control populations for COVID-19 (left), including protein targets with strong evidence for statistical colocalization for at least one definition (right). The column in the middle reports p-values. BGAT = Histo-blood group ABO system transferase; HSP 40 = DnaJ homolog subfamily B member 1; NUDT5 = ADP-sugar pyrophosphatase; RAB2A = Ras-related protein Rab-2A; ELF5 = ETS-related transcription factor Elf-5; OAS1 = 2’-5’-oligoadenylate synthase 1; SFTPD = Pulmonary surfactant-associated protein D; G-CSF = Granulocyte colony-stimulating factor

The remaining candidate proteins showed robust evidence for selected COVID-19 outcomes in our initial analysis (**Fig. 2**), with G-CSF showing suggestive evidence (regional PP=64%) for a shared genetic signal with greater susceptibility to infection and more severe COVID-19 prognosis, using multi-trait colocalisation^31^ (**Supplementary Fig. 1**).

### ELF5 is the candidate causal gene at 11p13 for severe COVID-19

The lead cis-pQTL for ELF5, rs766826 (MAF=35.9%), is in strong linkage disequilibrium (LD; r^2^=0.81) with a recently identified variant rs61882275 associated with severe COVID-19 in an independent study using whole genome sequencing^32^. The causal gene remained unidentified, but the two closest candidate genes are *ELF5* and *CAT*, whose encoded protein products are both captured by our proteomic data. We identify *ELF5* as the causal gene at this locus through a cluster of colocalising phenotypes, including three different COVID-19 outcomes, *ELF5* expression in the lung, and ELF5 abundances in plasma (PP=93%), with rs766826 being the most likely (PP=99%) underlying causal variant (**Fig. 3**). We further tested for colocalisation of the cis-pQTL for ELF5 and gene expression across all GTEx tissues and identify colocalisation specific to expression in lung but not in other tissues, including tissues with high ELF5 expression such as breast, prostate, or salivary glands (**Supplementary Fig. 2**). This finding points towards a specific role of the major C-allele of rs766826 in increasing the expression of *ELF5* in the lung (beta=0.24, p-value=5.3×10^−15^) with subsequent higher risk for severe COVID-19 (odds ratio=1.11, p-value<5.0×10^−6^) and higher abundances of ELF5 in blood (beta=0.06, p-value<5.4×10^−6^), the latter likely via cell turnover or injury of lung tissue since ELF5 is not predicted to be actively secreted into blood^33^.

**Figure 3.**
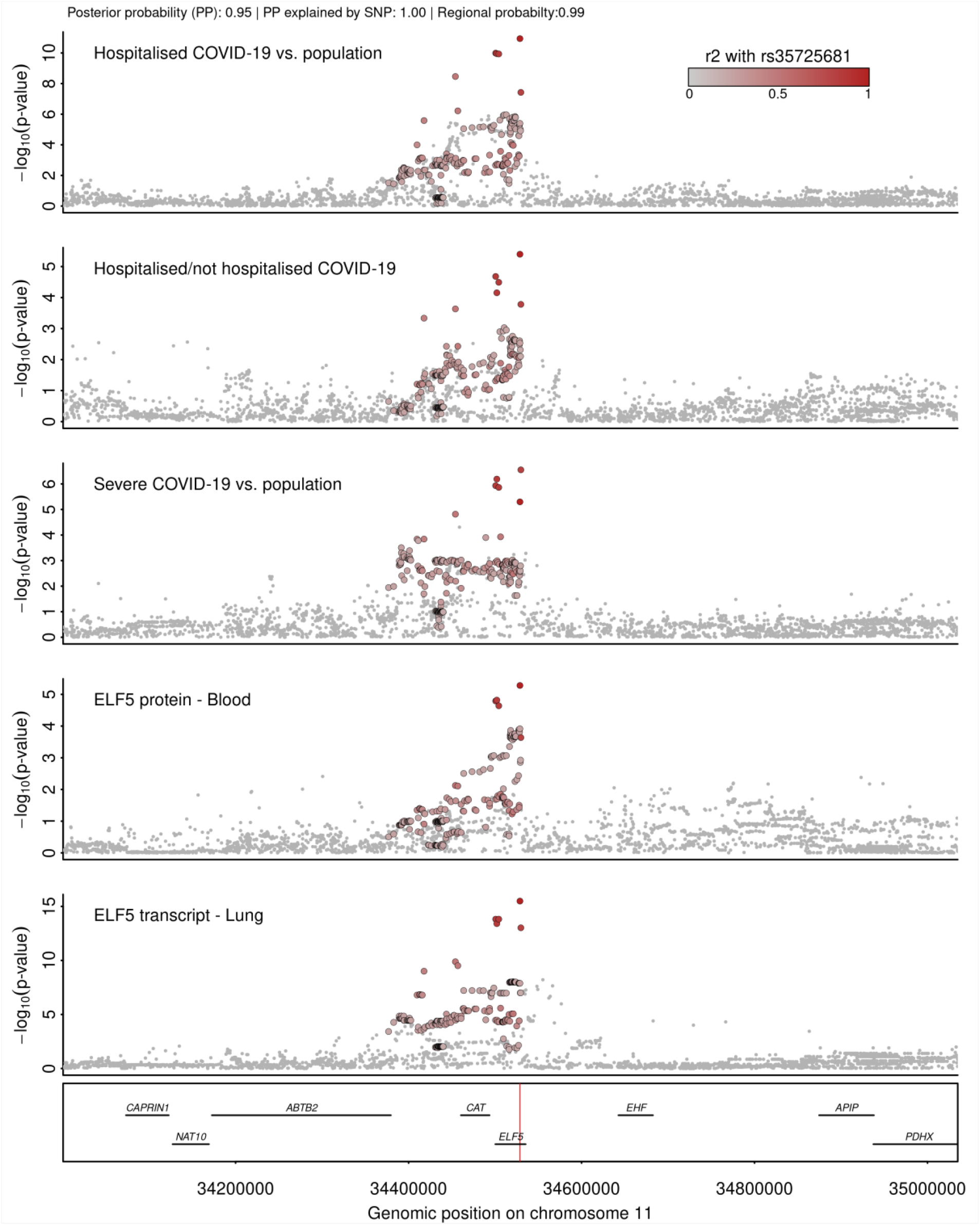
Stacked regional association plots at *ELF5*. Each panel contains regional association statistics (p-values) for the trait listed in the upper left corner along genomic coordinates. Each dots represents a single nucleotide polymorphisms and colours indicate linkage disequilibrium (LD; r^2^) with the most likely causal variant (rs766826) at this locus (darker colours stronger LD). The position of rs766826 in the genome is highlighted by a red line the lowest panel.

While we identified strong evidence for a shared genetic signal (PP>80%) between the lead cis-pQTL for the other candidate causal gene at this locus (*CAT*) and the corresponding cis-eQTL in 29 out of 49 tissues in GTEx v8, indicating convergence of gene and protein expression, plasma levels of catalase (encoded by *CAT*) were unrelated to COVID-19 phenotypes. We further observed that *CAT* expression in lung and whole blood formed a separate cluster (PP=56.6%) at the same genetic locus possibly explained by rs35725681 (PP=36.8%, **Supplementary Fig. 3**).

### rs766826 resides in an open chromatin region in the lung and is associated with lung function

While *ELF5* is known to be highly expressed in multiple tissues, we demonstrate that the genetic signal shared between ELF5, severe COVID-19 prognosis, and gene expression was specific to the lung. We observed that rs766826 mapped to an open chromatin region in AT2 cells from the lung but not in other tissues with high expression of *ELF5*, such as mammary gland, prostate, or kidney (**Supplementary Fig. 4**), providing a potential explanation for the tissue-specific effect.

To systematically test phenotypic consequences of rs766826 or proxies in strong LD (r^2^>0.8), we queried the OpenGWAS database^34^ and performed colocalization for all suggestive associations observed at p<10^−4^. The only association with evidence for a shared genetic signal with ELF5 expression and COVID-19 prognosis was seen for lung function (PP=99%) (**Supplementary Fig. 5**) with a somewhat counterintuitive effect direction seen for the ELF5-increasing and COVID-19 risk C-allele being associated with better lung function (beta=0.01, p-value<1.6×10^−5^) based on the quotient between forced expired volume in 1 second (FEV_1_) and forced vital capacity (FVC) from spirometry in population-based, that is, COVID-19 free studies^35^.

### ELF5 is expressed in epithelial cells of the respiratory system

We observed that *ELF5* was almost exclusively expressed by different epithelial cells of the respiratory system (**Fig. 4**) using single cell and single nucleus RNA sequencing (scnRNAseq) across different data sets of samples from healthy donors (**Fig. 4**, see **Methods**). Its expression pattern was shared with the viral entry receptor *ACE2* and associated proteases, such as *TMPRSS2*. Specifically, sustentacular and Bowman gland cells from the olfactory mucosa and secretory epithelial cells from the pseudostratified epithelium of the nasopharynx and distal bronchioles of the lung showed high expression levels of *ELF5* (**Fig. 4c**). They similarly expressed host proteins utilized by SARS-CoV-2, including *ACE2* or *TMPRSS2*, suggesting that putative target cells of SARS-CoV-2 express high quantities of *ELF5*.

Lungs from deceased COVID-19 patients consistently show signs of massive alveolar damage^16,36^. While *ELF5* expression was highest in secretory cells, AT2 but not AT1 cells showed consistent expression of *ELF5* in our lung data set of COVID-19-free donors^37^ (**Fig. 4**). This finding coincided with classical lineage markers for AT2 cells such as *SFTPD*, which was also highly expressed in mitotic cells (**Fig. 4C**). We note that our proteogenomic screen prioritised *SFTPD* as a candidate gene for severe COVID-19 in line with other studies^30,32^, leaving the possibility of potential interactions between candidate mediators.

**Figure 4.**
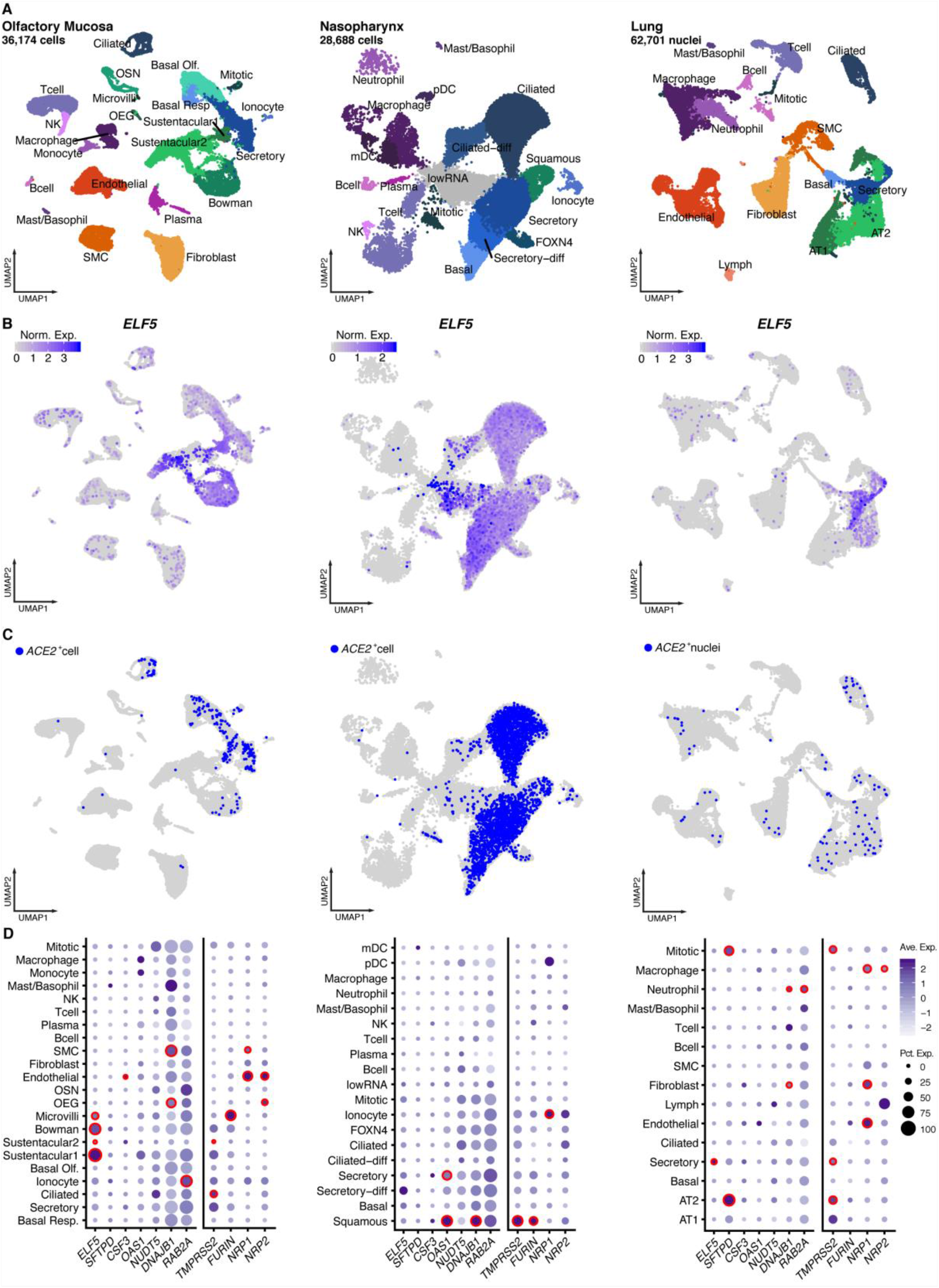
Expression of candidate genes in different single cell data sets covering the respiratory system. **A** UMAP representation of single cell/nuclei RNA sequencing data from three data sets (olfactory mucosa, nasopharynx, and lung) with annotations of cell types. **B** Expression levels of *ELF5* across all cells identified in all three data sets. **C** *ACE2*^+^ cells in all three data sets. **D** Dot plots showing the number of cells positive for candidate genes (size). The colour gradient indicates scaled average expression levels and red frames indicate significantly higher expression (adjusted p-value<0.05) of the target gene in one cell type compared to all others.

### Immunofluorescence staining validates scnRNAseq results and shows high ELF5 expression in AT2 cells of post-mortem COVID-19 samples

We validated the expression of ELF5 at the protein level in the different epithelial cells of the olfactory mucosa and lungs using immunofluorescence staining in non-COVID-19 samples (**Fig. 5** and **Supplementary Figure 6 and 7**). Within the olfactory mucosa, sustentacular cells (KRT18^+^) and horizontal basal cells (KRT18^-^) of the olfactory epithelium (above the dashed line, **Fig. 5A**) and the bowman gland cells (KRT18^+^) within the lamina propria (below the dashed line, **Fig. 5A**) were positively stained for ELF5 (**Fig. 5A** and **Supplementary Figure 6A**). We further validated protein expression of ELF5 in AT2 (SFTPC^+^) and epithelial cells (EPCAM^+^) of the airways in lung consistent with our scnRNAseq experiments (**Fig. 5B-C** and **Supplementary Figure 6B-D**). We observed similar validation of scnRNASeq experiments by immunofluorescence staining for ACE2 and TMPRSS2 (**Supplementary Figure 8**).

**Figure 5.**
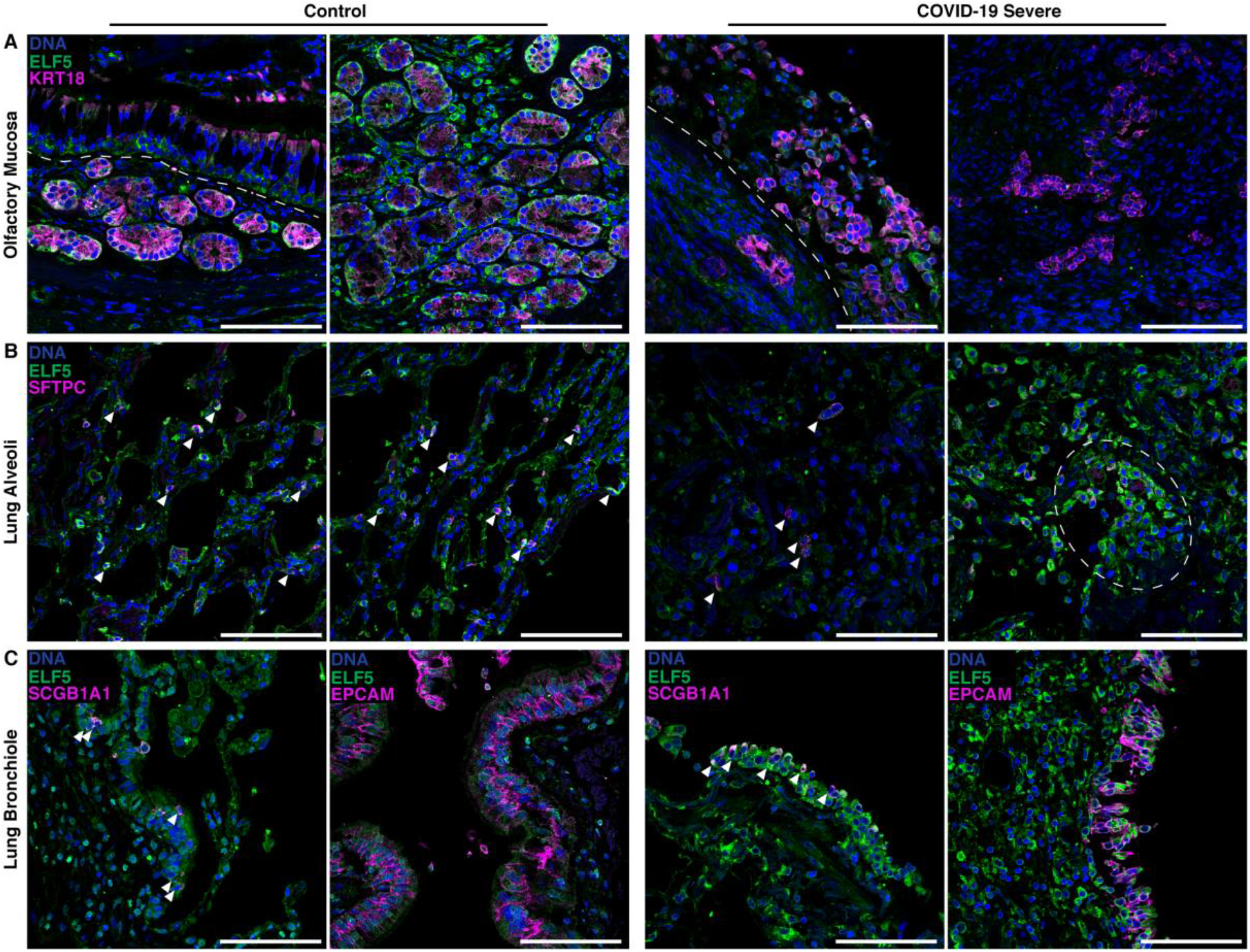
ELF5 expression by epithelial cells of the olfactory mucosa and lung. Immunofluorescent staining of ELF5 in control and COVID-19 patients in the **A** olfactory mucosa, **B** lung alveoli, and **C** lung bronchiole. **A** Dashed lines separate the olfactory epithelium and the lamina propria. **B** Arrowheads highlight AT2 cells expressing ELF5; dashed outline highlights clusters of AT cells expressing ELF5. **C** left: epithelial cells expressing ELF5; right: arrowheads highlight airway epithelial cells expressing ELF5. Marker genes for sustentacular and Bowman gland cells (**A**, KRT18), alveoli type II cells (**B**, SFTPC), pan-epithelial cells (**C**, EPCAM), and secretory cells (**C**, SCGB1A1) are shown in purple. Validation staining for each tissue: control (n=2); COVID-19 (n=2). Scale bar = 100μm

We next investigated ELF5 expression within the same tissues in samples from two patients who rapidly died from severe COVID-19 within ≤14 days (**Fig. 5A-C**). We observed an injured olfactory epithelium, including highly disrupted Bowman glands and very few cells showing ELF5 expression (**Fig. 5A**, left; **Supplementary Figure 6A**), and further loss of structural integrity of the alveolar region of the lung with only very few AT2 cells within the damaged region. However, AT2 cells characterized by high ELF5 expression also formed clusters, possibly reflective of their activated state to regenerate the epithelia^38,39^ (**Fig. 5B** and **Supplementary Figure 6B**). Similarly, secretory cells (SCGB1A1^+^) along with other epithelial cells of the airway mucosa (e.g., ciliated and basal cells) expressed ELF5 and were also injured over the course of SARS-CoV-2 infection (**Fig. 5C, Supplementary Figure 6C-D**). These observations suggest that *ELF5* expression might play a dynamic role during SARS-CoV-2 infection and COVID-19.

We finally observed potential signs of remodeling and high ELF5 expression in post-mortem respiratory tissue samples of two COVID-19 patients with a fatal but not rapid disease course due to intensive treatment, including extracorporeal membrane oxygenation (≥14 days, termed as ‘later death’; **Supplementary Figure 9A**). Mucosal structures were more similar to controls, although structural integrity was not fully restored. We further observed potentially regenerative structures with either AT2 cells or airway epithelial cells highly expressing ELF5 that may indicate an active wound healing response^39–41^ (**Supplementary Figure 9B-C**). Delorey *et al*. recently showed the induction of a regenerative program in cells of the airway and alveolar epithelium after SARS-CoV-2 infection^16^. However, AT2 cell renewal and AT1 cell differentiation were inhibited in COVID-19, leading to an accumulation of cells in this regenerative transitional cell state and potentially lung failure^16^.

### ELF5 and TMPRSS2 are co-expressed

To derive possible hypothesis of how *ELF5* expression might be linked to severe COVID-19, we collated a list of candidate genes, that were either regulated or co-expressed with *ELF5* (see **Methods** and **Supplementary Table 3**). Among the set of candidate genes were multiple members of the transmembrane serine protease-family, including *TMPRSS2* and *TMPRSS4*, which have been shown to be essential for viral entry by priming of the spike protein^42,43^. We observed a positive correlation between *ELF5* and *TMPRSS2* expression in sustentacular cells (r=0.15, p<4.5×10^−6^, **Supplementary Fig. 10**), the cell type with highest *ELF5* expression in our data, but no correlation with *TMPRSS4* expression. The correlation with *TMPRSS2* expression was also above the 95^th^ percentile of correlation coefficients across all genes. Further, genes highly correlated with *ELF5* expression were also significantly enriched among collated target genes, minimizing the possibility of a measurement artefact. While such correlation analysis using single-cell data must be treated with caution, overexpression of *Efl5* in a mouse model showed a three-fold increase in *Ace2* and a two-fold higher expression of *Tmprss4* in AT2 cells^44^, providing additional evidence that *ELF5* might be involved in the regulation of key host factors for SARS-CoV-2.

To formally test for pathways associated with *ELF5* expression, we performed cell-type specific gene set enrichment analysis using the collated set of putative ELF5 targets or co-expressed genes and observed a consistent enrichment of biosynthetic pathways like mRNA and peptide processing and possibly among genes involved in epithelial barrier formation (**Supplementary Fig. 11**). The latter aligns with a substantial impact of *Elf5* overexpression on the differentiation of the lung epithelium in mouse models of lung development, leading to dilation of the airways^44^.

### Drug target identification

We queried all identified candidate genes in the Open Targets database^45^ to identify repurposing opportunities for COVID-19. While none of the genes had already approved drugs or drugs in clinical trials, recombinant human G-CSF (rhG-CSF), such as filgrastim and lenograstim, is used to treat neutropenia caused by chemotherapy to stimulate production of granulocytes from the bone marrow^46^. In line with this, phenotypic associations identified in a phenome-wide analysis of the lead cis-pQTL for G-CSF showed that genetically higher plasma G-CSF was positively associated with granulocyte and other white blood cell counts (**Fig. 6 and Supplementary Tab. 4**). This highlights the ability of the cis-pQTL to instrument the function of the protein and allowed testing of the potential effect of G-CSF supplementation for (severe) COVID-19 *in silico*. We observed a 25% (odds ratio: 0.75; 95%-CI: 0.65-0.87; p-value<1.5×10^−4^) reduction in the risk for severe COVID-19 per 1 s.d. higher genetically predicted G-CSF (**Supplementary Tab. 2**). A previous smaller, independent study that used a different proteomic technology observed directionally consistent results, but did not reach statistical significance^47^. Together, these results provide *in silico* evidence that people with genetically higher plasma G-CSF abundances are less likely to develop severe COVID-19 and suggests that treatment with rhG-CSF might decrease the risk for symptomatic or even severe COVID-19. A recent randomized clinical trial^48^ among 200 COVID-19 patients with pneumonia and severe lymphopenia observed a significantly lower number of patients developing critical illness when treated within the first three days of inclusion with 5μg/kg rhG-CSF. However, leucocytosis was common in the treatment arm, including severe cases, which may limit the general application. For example, it might be conceivable that rhG-CSF treatment in COVID-19 patients with a strong immune response stimulates an adverse hyperinflammatory state and hence only a subgroup of COVID-19 patients might benefit.

**Figure 6.**
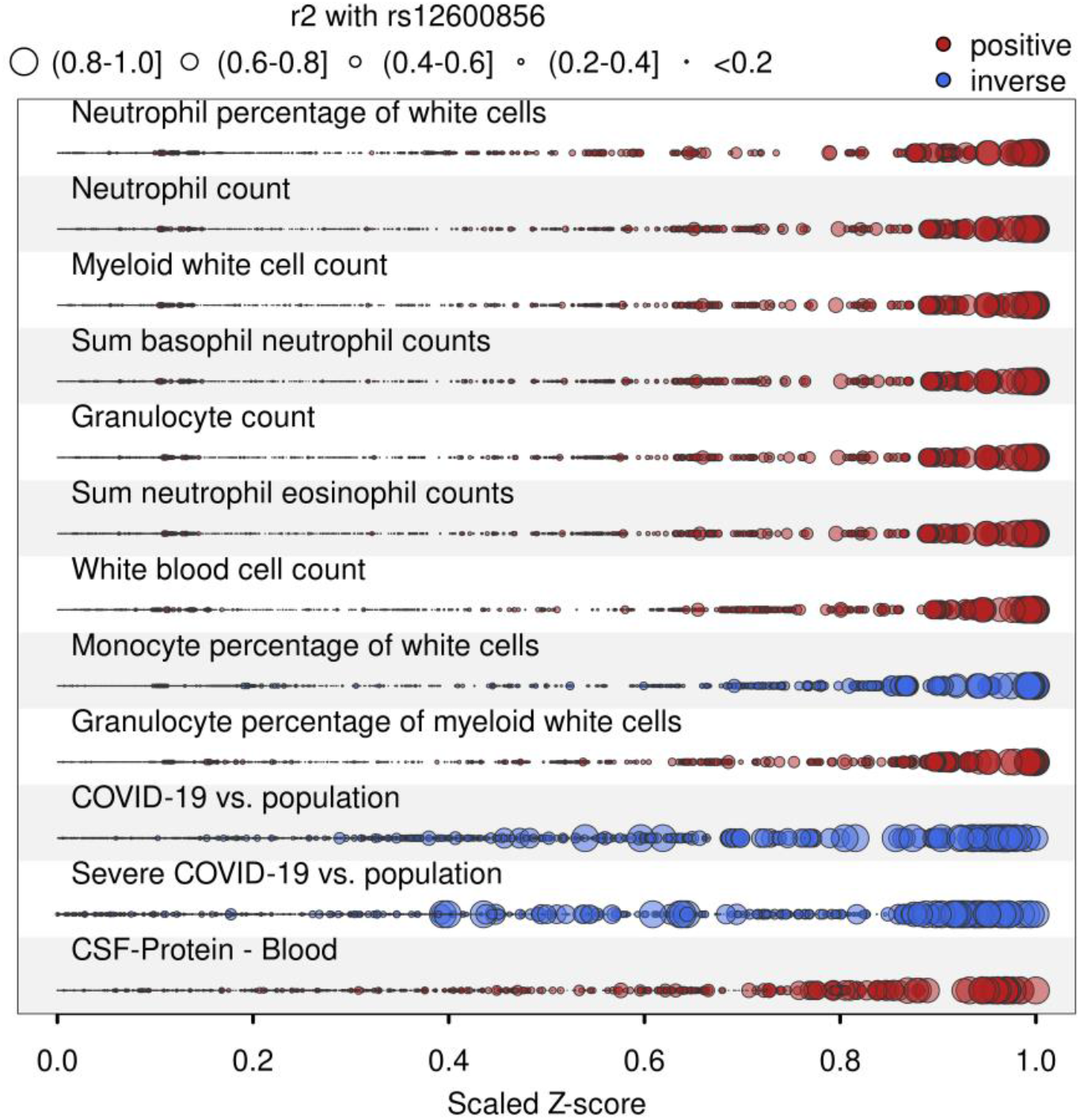
Convergence of genotype – phenotype variation at *GCSF3*. Plot visualizing convergence of genetic variants at the *CSF3* locus in relation to the linkage disequilibrium (LD) with the candidate gene variant identified by multi-trait colocalisation (rs12600856). Z-scores from genome-wide association studies for each annotated trait have been scaled by the absolute maximum, and dot size is proportional to the LD (r2). Colours indicate the direction of effect aligned to the risk-increasing allele (red – positive, blue - inverse). Effect estimates have been aligned to the protein increasing allele. Summary statistics for blood cell traits were obtained from Astle et al.^49^, statistics for COVID-19 and protein abundances as described in the main text.

## DISCUSSION

The identification of causal mechanisms that predispose infected individuals to a severe course of COVID-19, including hospitalisation and risk of death, can guide clinical management and the identification of novel drug targets or repurposing opportunities. We identified six not previously reported and replicated two known causal genes and their protein targets. We demonstrate that the strongest new and most robust candidate, *ELF5*, is specifically expressed in primary target cells of SARS-CoV-2 (for example, sustentacular^50^, AT2^51^, and secretory or ciliated epithelial cells^52^) with evidence of co-expression with genes encoding key host factors, such as *ACE2* and *TMPRSS2*, using scnRNAseq data across various sites of the respiratory system. We refine the association of genetic variation at *ELF5* with severe COVID-19 to a single causal variant (rs766826) with high confidence (PP=99%) and identify a tissue-specific effect of *ELF5* expression in lung that is associated with a more than 4-fold higher risk to develop severe COVID-19. Our results highlight high *ELF5* expression in primary target cells of SARS-CoV-2 as a possible mechanism for severe COVID-19 with a particular focus on epithelial cells.

We identify a total of eight candidate proteins with robust evidence for at least one outcome, with Ets-related transcription factor Elf-5 (ELF5) being the most consistent not yet reported candidate associated with a more than 4-fold higher genetically predicted risk for severe COVID-19. We show that *ELF5* is specifically expressed in primary target cells of SARS-CoV-2 of the upper and lower respiratory system, including sustentacular cells of the olfactory mucosa, secretory epithelial cells of the nasopharynx, and AT2 cells in the lung using single cell/nuclei RNA sequencing and immunohistochemistry. We further find genetically anchored evidence that aligns with a recent clinical trial^48^ suggesting human recombinant granulocyte colony-stimulating factor (G-CSF) as a potential treatment option among patients with COVID-19 and severe lymphopenia to mitigate adverse outcomes.

*ELF5* is a member of the Ets transcription factor family and is best known for its possible role in breast or prostate cancer, tissues with high fractions of epithelial cells^53^, and less for its possible role in lung development^44,54^ and possibly cystic fibrosis^55^. Experimental models to study the role of ELF5 are difficult since *Elf5*^*-/-*^ mice are embryonic lethal. However, the recent development of transgenic mouse models^56^ and our scnRNAseq data provide strong evidence that *ELF5* is expressed in epithelial cells of the respiratory system of adult mice and humans. Early work in lung tissue cultures and mouse models described a dynamic expression pattern of *Elf5* during embryogenesis and lung branching, including almost complete downregulation in distal lung postnatally, while residual expression in proximal airways persisted^44,54^. Overexpression of *Elf5* during early but not late embryonal development (after E16.5) caused a severe cystic lung phenotype characterised by disrupted branching and a dilated airway epithelium^44^, characteristics that are also seen in autopsies of COVID-19 patients^36^. While such a drastic intervention in mouse models is not comparable to the subtle effect of a common genetic variant, the observation that key host factors for SARS-CoV-2 (*Ace2* and *Tmprss4*) are upregulated in *Elf5*-overexpressing AT2 cells partly aligns with our observations using scRNAseq data.

The role of *ELF5* in secretory and AT2 cells of the airway and alveolar epithelium, respectively, may have potential implications to the wound healing response. As cells with stem-like capacity, they are involved in the maintenance and repair of their respective cellular niches^38,57^. Thus, any surviving secretory and AT2 cells that drive the repopulation of the epithelium could potentially have aberrant repair programs mediated by *ELF5* and therefore possibly rs766826, if any, and an accumulation of AT2 cells in a regenerative transitional cell state has recently been suggested for COVID-19^16^.

Up to 60% of COVID-19 patients report transient anosmia^58^, which is one of the few infectious symptoms with high specificity^59^. The underlying aetiology, however, remains largely elusive. Direct infection and hence damage of olfactory sensory neurons by SARS-CoV-2 could be one obvious explanation. Viral particles have been shown to be present in neuronal cells of the olfactory mucosa possibly presenting a route for CNS infection^60^, however, the generally undetectable expression levels of *ACE2* in those cells makes them an unlikely primary target compared to, for example, epithelial cells^50^. Previous studies suggested that the loss of essential supporting cells, sustentacular cells, in the olfactory mucosa causes anosmia^50,61^. Sustentacular cells have been suggested as primary targets of SARS-CoV-2 based on high *ACE2* expression^50,51,62^, supported by *in vivo* models showing a high viral load and rapid desquamation of the olfactory epithelium following infection^63,64^. A finding in line with our observations from samples of COVID-19 patients. Our observation that sustentacular cells, as well as other secretory epithelial cells in the olfactory mucosa, express high levels of *ELF5* along with a possible link to *ACE2* expression might indicate a possible modulating role of *ELF5* expression for this common symptom. However, a recent GWAS for anosmia^65^ among self-reported COVID-19 cases did not yet identify rs766826 and hence *ELF5* expression. While this might be explained by limited statistical power (the authors identified only one signal), it highlights that *ELF5* is likely only one out of many host factors possibly contributing to the onset of anosmia during SARS-CoV-2 infection.

We provide genetically anchored evidence that people with higher plasma G-CSF abundances are less likely to develop severe COVID-19, suggesting a possible protective effect possibly via early recruitment of neutrophils to the entry sites of SARS-CoV-2^66^. Colony-stimulating factors, such as G-CSF, are haematopoietic growth factors and are actively investigated as treatment options for COVID-19^67^. A recent open-label, multicentre, randomized clinical trial^48^ evaluated the efficacy of rhG-CSF to improve symptoms among 200 COVID-19 patients with lymphopenia (lymphocyte cell count <800 per μL) but without comorbidities. While no significant effect on the primary endpoint (time to improvement) was detected, patients treated with rhG-CSF experienced significantly fewer severe adverse effects, including respiratory failure, acute respiratory distress symptom, sepsis, or septic schock^48^. The treatment effect seemed further dependent on baseline lymphocyte counts, with patients <400 per μL benefiting the most. However, leucocytosis was common in the treatment arm, including severe cases. We note, that our results and the trial are in stark contrast to observational studies associating higher G-CSF plasma levels^68,69^ and rhG-CSF treatment among cancer patients with a poor prognosis^70,71^, likely explained by the inability to distinguish cause and effect. We further emphasize, that our genetically anchored drug prioritisation approach cannot make any recommendations about the best timepoint and dose of intervention during the course of infection/disease, which are crucial parameters for any drug application. Bespoke large randomized clinical trials are warranted to evaluate optimal timing, dosage, and risk-benefit evaluation of rhG-CSF treatment among COVID-19 patients.

Although the GWAS summary statistics from the COVID-19 Host Genetics Initiative represent multiple ancestries and the signal at the *ELF5* locus has recently been replicated in a Brazilian cohort^72^, the pQTL instruments are based on a single ancestry and genetic studies of plasma abundances of proteins in other ancestries may reveal additional candidate proteins, that may help to explain the variable prevalence of adverse COVID-19 outcomes across ethnicities^8^. We obtained some evidence that rs766826 might act through a mechanism that is possibly unique to AT2 cells based on an open chromatin region, the concrete underlying mechanism, however, remains elusive. Further studies are needed to decipher the role of rs766826 in the cell-type specific expression of *ELF5*. The same holds true for the suggested mechanisms of action for *ELF5*, for example co-expression with and possibly regulation of *ACE2* or *TMPRSS2*, that need to be tested in appropriate cellular and animal models, also to investigate the role of *ELF5* in tissues of the respiratory system more in general. Although our results started with the investigation of proteins measured in plasma and might hence provide possible biomarkers for severe COVID-19 in a clinical setting, we did not identify concordant associations based on plasma proteomic profiling for most of the candidates in public data sets^29,73^. This likely reflects a general segregation of proteins that possibly cause a more severe outcome of COVID-19 from those being a consequence of SARS-CoV-2 infection and COVID-19. We note that while MR can indicate direction of effects, estimates should be interpreted with caution when plasma/blood is not the tissue of action of the protein or if cis-pQTL(s) can be linked to protein altering variants or splicing event QTLs^74^, which likely explains the small effect sizes for BGAT (linked to a splicing QTL) or SFTPD (the cis-pQTL, rs721917, being a missense variant, p.M31T).

Our results demonstrate potential modulators for a poor prognosis among COVID-19 patients with potential therapeutic options. We identify *ELF5* as a potential regulator in cells that are the primary targets of SARS-CoV-2 by combining population-level genetic evidence with gene expression at single-cell resolution, providing tangible hypothesis for further functional follow-up studies to investigate the role of *ELF5* for viral entry and wound healing of the epithelial layer of the respiratory system upon severe COVID-19.

## METHODS and MATERIALS

### Summary statistics for proteins

We obtained locus-specific, ±500kB of the protein encoding gene, summary statistics from our genome-wide association analysis for 4,775 plasma proteins targeted by the SomaScan v4 assay and 1,069 targeted by the Olink proximity extension assay^28,75^. A detailed description can be found elsewhere^28,75^. Briefly, plasma abundances of 4,775 protein targets were tested for protein quantitative trait loci using standard GWAS workflows based on 10.4 million single nucleotide polymorphisms among 10,708 individuals of white British descent. We further obtained genome-wide association statistics for 1,069 proteins measured using the complementary Olink technique available among a subcohort of 485 participants. We tested all protein targets with at least suggestive evidence (p<10^−5^) of a cis-pQTL or a COVID-19 signal close to the protein-encoding gene. We further treated protein assays as separate instances even if those targeted the same protein between both techniques, given the heterogeneity of genetic findings across both platforms^74^. We tested a total of 2,375 protein targets (n=723 common to both platforms) for colocalisation with any of the four COVID-19 outcome definitions.

### Summary statistics for COVID-19

We used four meta-analysed COVID-19 data sets from the June 2021 release of the COVID-19 Host Genetics Initiative (https://www.covid19hg.org/results/r6/), comprising A2 (very severe respiratory confirmed COVID-19 vs. population), B1 (hospitalised COVID-19 vs. not hospitalised COVID-19), B2 (hospitalised COVID-19 vs. population), and C2 (COVID-19 vs. population). A summary of case definitions can be found in Supplementary Table 1.

### Statistical colocalisation

To identify proteins that share a genetic architecture with any of the four different COVID-19 outcomes we performed statistical colocalisation^76^ in a 500kb window around the protein coding gene as implemented in the R package *coloc*. Briefly, statistical colocalisation is a Bayesian approach that provides posterior probabilities for each of five hypothesis: H0 – none of two traits has a genetic signal in the region; H1 – only trait 1 has evidence for a genetic signal in the region; H2 – only trait 2 has evidence for a genetic signal in the region; H3 – both traits have two distinct signals in the same genomic region; and H4 – both traits share the same underlying genetic signal. We used default priors and further restricted colocalisation analysis to regions with at least suggestive evidence for a cis-pQTL (p<10^−5^). To accommodate the single variant assumption of *coloc*, we further required each protein-outcome pair passing the PP threshold of 80% and that respective regional lead variants are in strong LD (r^2^>0.8). To further acknowledge the fact that some cis-pQTLs are likely driven by measurement artefacts introduced by common but benign missense variation, we performed another round of colocalisation conditioning on the lead cis-pQTL in the region, if we had such evidence.

### Mendelian randomization

To derive effect directions and estimate possible effects of life-long higher/lower protein abundances on COVID-19 susceptibility and severeness, we performed single-instrument Mendelian randomization (MR) analysis using cis protein quantitative trait loci (cis-pQTLs) as instruments. We computed the Wald ratio^77^ to derive an estimate for the causal effect of a 1 s.d. increase in plasma abundances of the candidate protein on the risk for COVID-19.

### Tissue gene expression

We incorporated gene expression data by testing for a shared genetic signal between protein abundance in plasma and expression of the protein encoding gene in one of at least 49 tissues of the GTX v8 resource^78^. We used the same colocalisation approach as described above.

### Phenotypic follow-up of candidate cis-pQTLs

We systematically tested for phenotypic associations for cis-pQTLs by querying the OpenGWAS^34^ database, including proxies in high LD (r^2^>0.8). To test for a shared genetic signal between the FEV1/FCV ratio (a proxy for lung function) and plasma levels of ELF5, we downloaded genome-wide summary statistics from Shrine et al.^35^. We conditioned on two stronger independent lead signals in the region (rs10836366 and rs1648123) to account for the single variant assumption in statistical colocalisation.

### Collation of target genes of ELF5

We collated a list of genes with possible direction association with *ELF5* by querying the Molecular Signatures Data Base^79^, the Enricr tool^80^, the Harmonizome^81^, including ChIP-Seq experiments^82^, and a curated gene co-expression network^83^ (**Supplementary Table 3**).

### Single-cell/nucleus RNA sequencing

Single-cell/nucleus RNA sequencing (sc/nRNA-seq) healthy control datasets of the olfactory mucosa (GSE139522), nasopharynx (EGAS00001005461), and lungs (EGAS00001004689; EGAS00001004419: SAMEA6848756, SAMEA6848761, SAMEA6848765, SAMEA6848766) were reanalyzed in this study^37,66,84,85^. Due to the increased noise of snRNA-seq data, we performed ambient RNA removal on the lung dataset with SoupX v1.4.5^86^. Analysis was performed with Seurat v3.1.4^87,88^. For the olfactory mucosa and lung datasets, individual samples were integrated and annotated from scratch. Individual samples were subjected to a upper bound filter of <10% mitochondrial reads and >200 genes expressed, and an upper bound filter of 3000-6000 genes depending on the sample. After log-normalization and scaling, canonical correlation analysis was used for integration and batch-correction of the individual samples. Principal component analysis and Uniform Manifold Approximation and Projection for dimension reduction (UMAP) were calculated for each integrated dataset. Finally, after unsupervised clustering, cell type assignment was performed as previously described^15,37,50,66,84,85^ and marker genes are depicted in **Supplementary Figure 11**. Differentially expressed genes were identified using a MAST-based differential expression test. The Pearson’s correlation was calculated on log-normalized expression values for all detected genes against *ELF5* in Sustentacular1 cells, which expressed *ELF5* the highest. Gene set enrichment analysis^79^ (GSEA; v4.1.0) was used to test for enrichment of the collated *ELF5* target genes against all detected genes where the weights used were the Pearson’s correlation values. Utilizing the collated *ELF5* target genes, an *ELF5* target gene expression score was calculated for all cells using the AdddModuleScore() from Seurat.

### Immunohistochemistry

Postmortem olfactory mucosa and lung tissue were collected from control and COVID-19 donors. Non-COVID-19 samples were obtained from the BrainBank/Biobank of the Department of Neuropathology at the Charité – Universitätsmedizin Berlin (**Supplementary Tab. 5**). COVID-19 status from the BrainBank/Biobank samples was assessed by Spindiag Rhonda PCR rapid COVID-19 test according to the manufactures protocol. This study was approved by the local ethics committees (EA1/144/13, EA2/066/20 and EA1/075/19) as well as by the Charité–BIH COVID-19 research board and was in compliance with the Declaration of Helsinki; autopsies were performed on the legal basis of §1 of the Autopsy Act of the state Berlin and §25(4) of the German Infection Protection Act. Control lung tissues were purchased from OriGene (TissueFocus) and Tissue Solutions. Samples were embedded in paraffin and sectioned at a 5μm thickness. Sections were deparaffinized in Roticlear (CarlRoth, A538.1) and rehydrated with an ethanol series. Antigen retrieval was performed by submerging slides in 10mM Sodium Citrate Buffer (Sigma-Aldrich, C999-1000ML) at 95°C for 10 minutes and left to cool for 30 minutes. Sections were permeabilized and blocked with 5% goat serum PBST (0.5% Triton X-100 in PBS) for 1 hour. Primary antibodies (1% goat serum PBST) were then added onto the tissues and left to incubate overnight. This was followed by secondary antibody (1% goat serum PBST) labelling for 1 hour at room temperature. Antibodies used in this study can be found in **Supplementary Table 6**. TMPRSS2 with ACE2 staining was performed according to the instructions of VectaFluor™ Excel Amplified Kit (Vector Laboratories; DK-2594). To remove autofluorescence, sections were sequentially treated with Lipofuscin Autofluorescence Quencher (PromoCell, PK-CA707-23007) and Vector TrueVIEW™ Kit (Vector Laboratories, SP-8400). Sections were then stained with 16μM Hoechst 33258 (ThermoFischer, H3569) for 5 minutes, washed with 1X PBS, and mounted with VECTASHIELD® HardSet™ Antifade Mounting Medium (Vector Laboratories, H-1400-10). Stained slides were visualized with a Leica SP8 confocal microscope. Images were processed and assembled with FIJI^89^.

## Data Availability

Summary statistics for protein levels are available from www.omicscience.org. Summary statistics for COVID–19 are available from https://www.covid19hg.org/results/r6/. scRNAseq data sets are available under the accession IDs: olfactory mucosa (GSE139522), nasopharynx (EGAS00001005461), and lungs (EGAS00001004689, EGAS00001004419, SAMEA6848756, SAMEA6848761, SAMEA6848765, SAMEA6848766).

https://www.covid19hg.org/results/r6/

## ACKNOWLEDGEMENTS

We are grateful to all Fenland volunteers and to the General Practitioners and practice staff for assistance with recruitment. We thank the Fenland Study Investigators, Fenland Study Co-ordination team and the Epidemiology Field, Data and Laboratory teams. Proteomic measurements were supported and governed by a collaboration agreement between the University of Cambridge and SomaLogic.

## FUNDING

The Fenland Study (10.22025/2017.10.101.00001) is funded by the Medical Research Council (MC_UU_12015/1). We further acknowledge support for genomics from the Medical Research Council (MC_PC_13046). CL, EW, MP, and NJW are funded by the Medical Research Council (MC_UU_00006/1 - Aetiology and Mechanisms). Non-COVID-19 autopsy samples were provided by the BrainBank/BioBank of the Department of Neuropathology at the Charité – Universitätsmedizin Berlin, funded by the Deutsche Forschungsgemeinschaft (DFG, German Research Foundation) under Germany’s Excellence Strategy – EXC-2049 – 390688087. COVID-19 autopsies were supported by the the German Network University Medicine NUM FKZ 01KX2021 Organostrat and Defeat Pandemics (H.R. and F.L.H.).

## AUTHOR CONTRIBUTIONS

Conceptualization: CL, MP, CC

Data curation/Software: MP, RLC, SL, EW

Formal Analysis: MP, RLC, SL, EW

Methodology: SL, ST

Visualization: MP, RLC, SL, KJ

Experiments: RLC, KJ, HR

Funding acquisition: CL, NJW, CC, IL, RE

Project administration: CL, NJW, CC, RE, IL

Supervision: CL, CC

Writing – original draft: MP, RCL, SL, CC, CL

Writing – review & editing: EW, HR, ST, BH, RE, MM, LS, IL, NJW

## COMPETING INTERESTS

All other authors declare that they have no competing interests.

## DATA and MATERIALS AVAILABILITY

Summary statistics for protein levels are available from www.omicscience.org. Summary statistics for COVID-19 are available from https://www.covid19hg.org/results/r6/. scRNAseq data sets are available under the accession IDs: olfactory mucosa (GSE139522), nasopharynx (EGAS00001005461), and lungs (EGAS00001004689, EGAS00001004419, SAMEA6848756, SAMEA6848761, SAMEA6848765, SAMEA6848766). Associated code and scripts for the analysis will be made available on GitHub upon publication (https://github.com/pietznerm/elf5_covid19).

## Notes

### Competing Interest Statement

The authors have declared no competing interest.

### Author Declarations

This study was approved by the local ethics committees (Berlin: EA1/144/13, EA2/066/20 and EA1/075/19) as well as by the Charite–BIH COVID–19 research board and was in compliance with the Declaration of Helsinki; autopsies were performed on the legal basis of paragraph 1 of the Autopsy Act of the state Berlin and paragraph 25(4) of the German Infection Protection Act.

## REFERENCES

1. Fauci, A. S. The story behind COVID-19 vaccines. Science 372, 109 (2021).

2. Haas, E. J. et al. Infections, hospitalisations, and deaths averted via a nationwide vaccination campaign using the Pfizer-BioNTech BNT162b2 mRNA COVID-19 vaccine in Israel: a retrospective surveillance study. Lancet. Infect. Dis. 3099, 1–10 (2021).

3. Dan, J. M. et al. Immunological memory to SARS-CoV-2 assessed for up to 8 months after infection. Science 371, (2021).

4. Planas, D. et al. Reduced sensitivity of SARS-CoV-2 variant Delta to antibody neutralization. Nature 596, 276–280 (2021).

5. Davies, N. G. et al. Estimated transmissibility and impact of SARS-CoV-2 lineage B.1.1.7 in England. Science 372, (2021).

6. Lemey, P. et al. Untangling introductions and persistence in COVID-19 resurgence in Europe. Nature 595, 713–717 (2021).

7. Wagner, C. E. et al. Vaccine nationalism and the dynamics and control of SARS-CoV-2. Science 373, eabj7364 (2021).

8. Williamson, E. J. et al. Factors associated with COVID-19-related death using OpenSAFELY. Nature 584, 430–436 (2020).

9. Grasselli, G. et al. Risk Factors Associated with Mortality among Patients with COVID-19 in Intensive Care Units in Lombardy, Italy. JAMA Intern. Med. 180, 1345–1355 (2020).

10. Pijls, B. G. et al. Demographic risk factors for COVID-19 infection, severity, ICU admission and death: a meta-analysis of 59 studies. BMJ Open 11, e044640 (2021).

11. Gao, Y.-D. et al. Risk factors for severe and critically ill COVID-19 patients: A review. Allergy 76, 428–455 (2021).

12. Pairo-Castineira, E. et al. Genetic mechanisms of critical illness in COVID-19. Nature 591, 92– 98 (2021).

13. Ellinghaus, D. et al. Genomewide Association Study of Severe Covid-19 with Respiratory Failure. N. Engl. J. Med. (2020) doi:10.1056/NEJMoa2020283.

14. Initiative, C.-19 H. G. Mapping the human genetic architecture of COVID-19. Nature (2021) doi:10.1038/s41586-021-03767-x.

15. Chua, R. L. et al. COVID-19 severity correlates with airway epithelium-immune cell interactions identified by single-cell analysis. Nat. Biotechnol. 38, 970–979 (2020).

16. Delorey, T. M. et al. COVID-19 tissue atlases reveal SARS-CoV-2 pathology and cellular targets. Nature 595, 107–113 (2021).

17. Melms, J. C. et al. A molecular single-cell lung atlas of lethal COVID-19. Nature 595, 114–119 (2021).

18. Karki, R. et al. Synergism of TNF-α and IFN-γ Triggers Inflammatory Cell Death, Tissue Damage, and Mortality in SARS-CoV-2 Infection and Cytokine Shock Syndromes. Cell 184, 149-168.e17 (2021).

19. Lee, S. et al. Virus-induced senescence is driver and therapeutic target in COVID-19. Nature (2021) doi:10.1038/s41586-021-03995-1.

20. Trump, S. et al. Hypertension delays viral clearance and exacerbates airway hyperinflammation in patients with COVID-19. Nat. Biotechnol. 39, 705–716 (2021).

21. Downes, D. J. et al. Identification of LZTFL1 as a candidate effector gene at a COVID-19 risk locus. Nat. Genet. (2021) doi:10.1038/s41588-021-00955-3.

22. Zhou, S. et al. A Neanderthal OAS1 isoform protects individuals of European ancestry against COVID-19 susceptibility and severity. Nat. Med. 27, 659–667 (2021).

23. Wickenhagen, A. et al. A prenylated dsRNA sensor protects against severe COVID-19. Science 3, eabj3624 (2021).

24. Gaziano, L. et al. Actionable druggable genome-wide Mendelian randomization identifies repurposing opportunities for COVID-19. Nat. Med. 27, 668–676 (2021).

25. Anisul, M. et al. A proteome-wide genetic investigation identifies several SARS-CoV-2-exploited host targets of clinical relevance. Elife 10, (2021).

26. Pathak, G. A. et al. Integrative genomic analyses identify susceptibility genes underlying COVID-19 hospitalization. Nat. Commun. 12, 4569 (2021).

27. Klaric, L. et al. Mendelian randomisation identifies alternative splicing of the FAS death receptor as a mediator of severe COVID-19. medRxiv Prepr. Serv. Heal. Sci. 1–28 (2021) doi:10.1101/2021.04.01.21254789.

28. Pietzner, M. et al. Mapping the proteo-genomic convergence of human diseases. Science 374, eabj1541 (2021).

29. Filbin, M. R. et al. Longitudinal proteomic analysis of severe COVID-19 reveals survival-associated signatures, tissue-specific cell death, and cell-cell interactions. Cell Reports Med. 2, (2021).

30. Initiative, C.-19 H. G. & Ganna, A. Mapping the human genetic architecture of COVID-19: an update. medRxiv 2021.11.08.21265944 (2021) doi:10.1101/2021.11.08.21265944.

31. Foley, C. N. et al. A fast and efficient colocalization algorithm for identifying shared genetic risk factors across multiple traits. Nat. Commun. 12, 764 (2021).

32. Kousathanas, A. et al. Whole genome sequencing identifies multiple loci for critical illness caused by COVID-19. medRxiv 2021.09.02.21262965 (2021) doi:10.1101/2021.09.02.21262965.

33. Uhlén, M. et al. The human secretome. 0274, 1–9 (2019).

34. Elsworth, B. et al. The MRC IEU OpenGWAS data infrastructure. bioRxiv 2020.08.10.244293 (2020) doi:10.1101/2020.08.10.244293.

35. Shrine, N. et al. New genetic signals for lung function highlight pathways and chronic obstructive pulmonary disease associations across multiple ancestries. Nat. Genet. 51, 481– 493 (2019).

36. Carsana, L. et al. Pulmonary post-mortem findings in a series of COVID-19 cases from northern Italy: a two-centre descriptive study. Lancet Infect. Dis. 20, 1135–1140 (2020).

37. Lukassen, S. et al. SARS -CoV-2 receptor ACE 2 and TMPRSS 2 are primarily expressed in bronchial transient secretory cells. EMBO J. 39, 1–15 (2020).

38. Barkauskas, C. E. et al. Type 2 alveolar cells are stem cells in adult lung. J. Clin. Invest. 123, 3025–36 (2013).

39. Desai, T. J., Brownfield, D. G. & Krasnow, M. A. Alveolar progenitor and stem cells in lung development, renewal and cancer. Nature 507, 190–4 (2014).

40. Tata, P. R. et al. Dedifferentiation of committed epithelial cells into stem cells in vivo. Nature 503, 218–23 (2013).

41. Rock, J. R. et al. Multiple stromal populations contribute to pulmonary fibrosis without evidence for epithelial to mesenchymal transition. Proc. Natl. Acad. Sci. U. S. A. 108, E1475–83 (2011).

42. Hoffmann, M. et al. SARS-CoV-2 Cell Entry Depends on ACE2 and TMPRSS2 and Is Blocked by a Clinically Proven Protease Inhibitor. Cell 181, 271-280.e8 (2020).

43. Zang, R. et al. TMPRSS2 and TMPRSS4 promote SARS-CoV-2 infection of human small intestinal enterocytes. Sci. Immunol vol. 5 https://www.science.org (2020).

44. Metzger, D. E., Stahlman, M. T. & Shannon, J. M. Misexpression of ELF5 disrupts lung branching and inhibits epithelial differentiation. Dev. Biol. 320, 149–160 (2008).

45. Ochoa, D. et al. Open Targets Platform: supporting systematic drug-target identification and prioritisation. Nucleic Acids Res. 49, D1302–D1310 (2021).

46. Gunzer, K. et al. Contribution of glycosylated recombinant human granulocyte colony-stimulating factor (lenograstim) use in current cancer treatment: review of clinical data. Expert Opin. Biol. Ther. 10, 615–630 (2010).

47. Li, M., Ho, C., Yeung, C. H. C. & Schooling, C. M. Circulating Cytokines and Coronavirus Disease: A Bi-Directional Mendelian Randomization Study. Front. Genet. 12, 680646 (2021).

48. Cheng, L.-L. et al. Effect of Recombinant Human Granulocyte Colony-Stimulating Factor for Patients With Coronavirus Disease 2019 (COVID-19) and Lymphopenia: A Randomized Clinical Trial. JAMA Intern. Med. 181, 71–78 (2021).

49. Astle, W. J. et al. The Allelic Landscape of Human Blood Cell Trait Variation and Links to Common Complex Disease. Cell 167, 1415-1429.e19 (2016).

50. Brann, D. H. et al. Non-neuronal expression of SARS-CoV-2 entry genes in the olfactory system suggests mechanisms underlying COVID-19-associated anosmia. Sci. Adv. 6, 5801–5832 (2020).

51. Muus, C. et al. Single-cell meta-analysis of SARS-CoV-2 entry genes across tissues and demographics. Nat. Med. 27, 546–559 (2021).

52. Hou, Y. J. et al. SARS-CoV-2 Reverse Genetics Reveals a Variable Infection Gradient in the Respiratory Tract. Cell 182, 429-446.e14 (2020).

53. Piggin, C. L. et al. ELF5 isoform expression is tissue-specific and significantly altered in cancer. Breast Cancer Res. 18, 1–18 (2016).

54. Metzger, D. E., Xu, Y. & Shannon, J. M. Elf5 Is an Epithelium-Specific, Fibroblast Growth Factor-Sensitive Transcription Factor in the Embryonic Lung. Dev. Dyn. 236, 1175–1192 (2007).

55. Swahn, H. et al. Coordinate regulation of ELF5 and EHF at the chr11p13 CF modifier region. J. Cell. Mol. Med. 23, 7726–7740 (2019).

56. Singh, S. et al. A new Elf5CreERT2-GFP BAC transgenic mouse model for tracing Elf5 cell lineages in adult tissues. FEBS Lett. 593, 1030–1039 (2019).

57. Rawlins, E. L. et al. The role of Scgb1a1+ Clara cells in the long-term maintenance and repair of lung airway, but not alveolar, epithelium. Cell Stem Cell 4, 525–34 (2009).

58. Menni, C. et al. Real-time tracking of self-reported symptoms to predict potential COVID-19. Nat. Med. 26, 1037–1040 (2020).

59. Struyf, T. et al. Signs and symptoms to determine if a patient presenting in primary care or hospital outpatient settings has COVID-19. Cochrane database Syst. Rev. 2, CD013665 (2021).

60. Meinhardt, J. et al. Olfactory transmucosal SARS-CoV-2 invasion as a port of central nervous system entry in individuals with COVID-19. Nat. Neurosci. 24, 168–175 (2021).

61. Khan, M. et al. Visualizing in deceased COVID-19 patients how SARS-CoV-2 attacks the respiratory and olfactory mucosae but spares the olfactory bulb. Cell 100264 (2021) doi:10.1016/j.cell.2021.10.027.

62. Fodoulian, L. et al. SARS-CoV-2 Receptors and Entry Genes Are Expressed in the Human Olfactory Neuroepithelium and Brain. iScience 23, 101839 (2020).

63. Bryche, B. et al. Massive transient damage of the olfactory epithelium associated with infection of sustentacular cells by SARS-CoV-2 in golden Syrian hamsters. Brain. Behav. Immun. 89, 579–586 (2020).

64. Ye, Q. et al. SARS-CoV-2 infection in the mouse olfactory system. Cell Discov. 7, 49 (2021).

65. Shelton, A. J. F., Shastri, A. J., Team, T. C.-& Aslibekyan, S. The UGT2A1 / UGT2A2 locus is associated with COVID-19-related anosmia. (2021).

66. Loske, J. et al. Pre-activated antiviral innate immunity in the upper airways controls early SARS-CoV-2 infection in children. Nat. Biotechnol. (2021) doi:10.1038/s41587-021-01037-9.

67. Lang, F. M., Lee, K. M. C., Teijaro, J. R., Becher, B. & Hamilton, J. A. GM-CSF-based treatments in COVID-19: reconciling opposing therapeutic approaches. Nat. Rev. Immunol. 20, 507–514 (2020).

68. Mudd, P. A. et al. Distinct inflammatory profiles distinguish COVID-19 from influenza with limited contributions from cytokine storm. Sci. Adv. 6, 16–18 (2020).

69. Meizlish, M. L. et al. A neutrophil activation signature predicts critical illness and mortality in COVID-19. Blood Adv. 5, 1164–1177 (2021).

70. Zhang, A. W. et al. The Effect of Neutropenia and Filgrastim (G-CSF) on Cancer Patients With Coronavirus Disease 2019 (COVID-19) Infection. Clin. Infect. Dis. 2019, 1–8 (2021).

71. Sereno, M. et al. A Multicenter Analysis of the Outcome of Cancer Patients with Neutropenia and COVID-19 Optionally Treated with Granulocyte-Colony Stimulating Factor (G-CSF): A Comparative Analysis. Cancers (Basel). 13, (2021).

72. Pereira, A. C. et al. Genetic risk factors and Covid-19 severity in Brazil: results from BRACOVID Study. medRxiv 2021.10.06.21264631 (2021) doi:10.1101/2021.10.06.21264631.

73. Overmyer, K. A. et al. Large-Scale Multi-omic Analysis of COVID-19 Severity. Cell Syst. 1–18 (2020) doi:10.1016/j.cels.2020.10.003.

74. Pietzner, M. et al. Cross-platform proteomics to advance genetic prioritisation strategies. bioRxiv 2021.03.18.435919 (2021) doi:10.1101/2021.03.18.435919.

75. Pietzner, M. et al. Synergistic insights into human health from aptamer-and antibody-based proteomic profiling. Nat. Commun. 12, 6822 (2021).

76. Giambartolomei, C. et al. Bayesian test for colocalisation between pairs of genetic association studies using summary statistics. PLoS Genet. 10, e1004383 (2014).

77. Lawlor, D. A., Harbord, R. M., Sterne, J. A. C., Timpson, N. & Davey Smith, G. Mendelian randomization: using genes as instruments for making causal inferences in epidemiology. Stat. Med. 27, 1133–63 (2008).

78. GTEx Consortium. The GTEx Consortium atlas of genetic regulatory effects across human tissues. Science 369, 1318–1330 (2020).

79. Subramanian, A. et al. Gene set enrichment analysis: a knowledge-based approach for interpreting genome-wide expression profiles. Proc. Natl. Acad. Sci. U. S. A. 102, 15545–50 (2005).

80. Kuleshov, M. V. et al. Enrichr: a comprehensive gene set enrichment analysis web server 2016 update. Nucleic Acids Res. 44, W90–7 (2016).

81. Rouillard, A. D. et al. The harmonizome: a collection of processed datasets gathered to serve and mine knowledge about genes and proteins. Database (Oxford). 2016, 1–16 (2016).

82. Kalyuga, M. et al. ELF5 suppresses estrogen sensitivity and underpins the acquisition of antiestrogen resistance in luminal breast cancer. PLoS Biol. 10, e1001461 (2012).

83. Deelen, P. et al. Improving the diagnostic yield of exome-sequencing by predicting gene-phenotype associations using large-scale gene expression analysis. Nat. Commun. 10, 2837 (2019).

84. Gassen, N. C. et al. SARS-CoV-2-mediated dysregulation of metabolism and autophagy uncovers host-targeting antivirals. Nat. Commun. 12, 3818 (2021).

85. Durante, M. A. et al. Single-cell analysis of olfactory neurogenesis and differentiation in adult humans. Nat. Neurosci. 23, 323–326 (2020).

86. Young, M. D. & Behjati, S. SoupX removes ambient RNA contamination from droplet-based single-cell RNA sequencing data. Gigascience 9, 1–10 (2020).

87. Butler, A., Hoffman, P., Smibert, P., Papalexi, E. & Satija, R. Integrating single-cell transcriptomic data across different conditions, technologies, and species. Nat. Biotechnol. 36, 411–420 (2018).

88. Stuart, T. et al. Comprehensive Integration of Single-Cell Data. Cell 177, 1888-1902.e21 (2019).

89. Schindelin, J. et al. Fiji: an open-source platform for biological-image analysis. Nat. Methods 9, 676–82 (2012).

