## Supplementary Figures 1-12 for "*ELF5* is a respiratory epithelial cell-specific risk gene for severe COVID-19"

**Affiliations**

### FIGURES

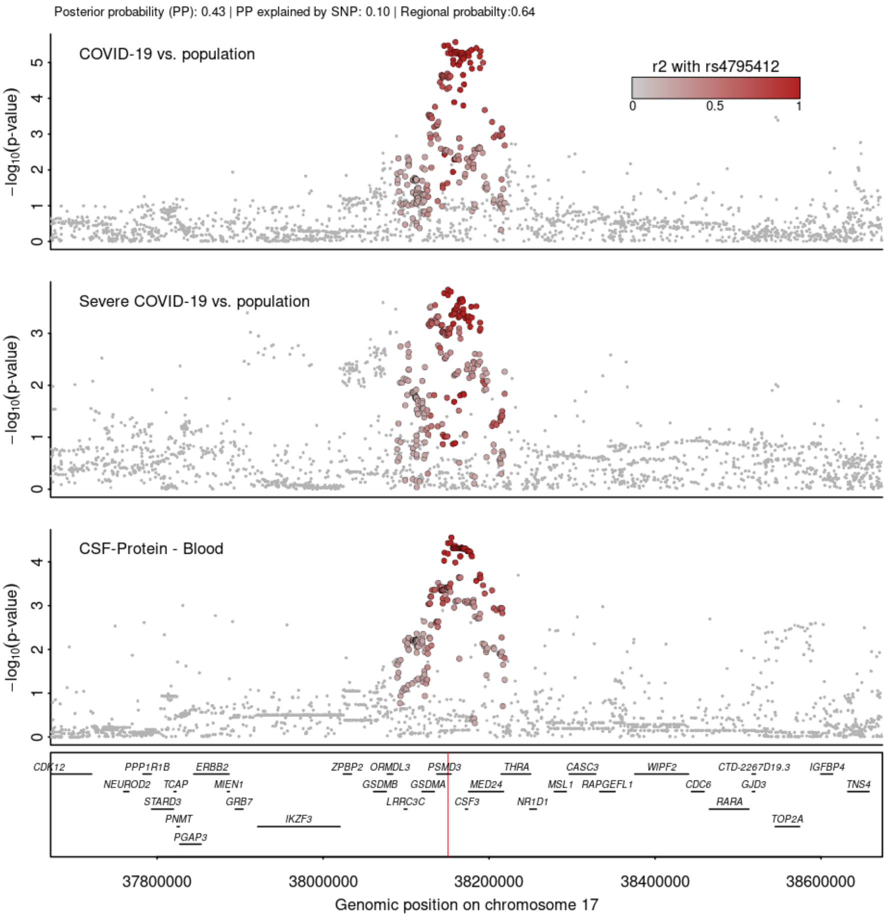


**Supplementary Figure 1 Stacked regional association plots at *CSF3*.** Each panel contains regional association statistics (p-values) for the trait listed in the upper left corner along genomic coordinates. Each dot represents a single nucleotide polymorphisms and colours indicate linkage disequilibrium (LD; $r^2$) with the most likely causative variant (rs4795412) at this locus (darker colours stronger LD).

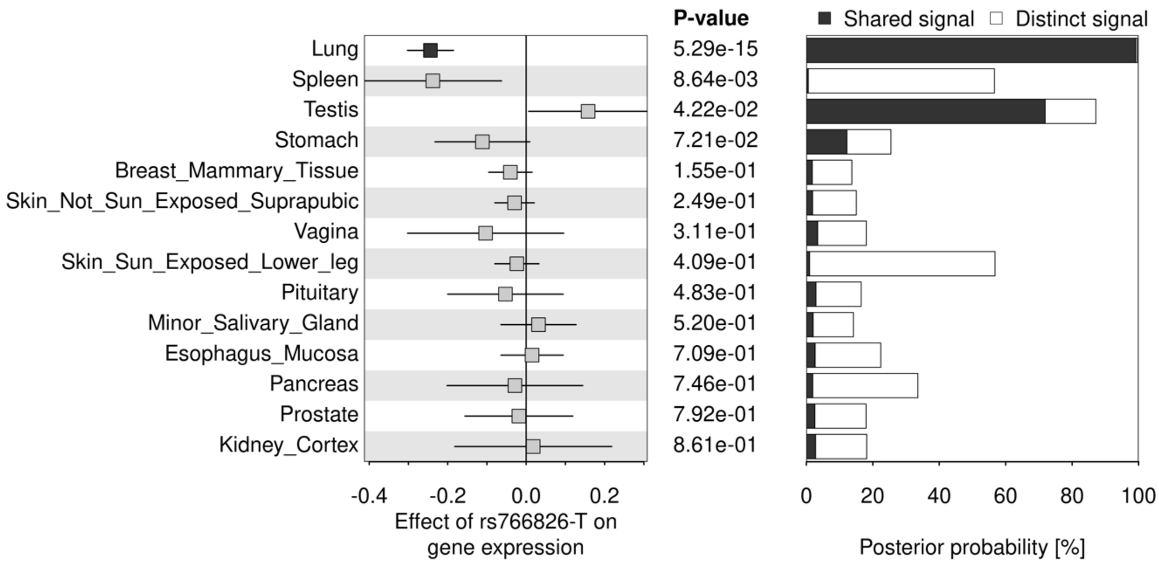


**Supplementary Figure 2** Summary of cross-tissue colocalisation for ELF5. The left panel shows effect estimates for rs766826 on *ELF5* expression in all tissues with detectable levels in the GTEx v8 resource. Significant effects (p<0.001) are highlighted in black. The left hand side shows posterior probabilities for testing for a shared genetic signal between ELF5 abundance in plasma and *ELF5* expression in each tissue.

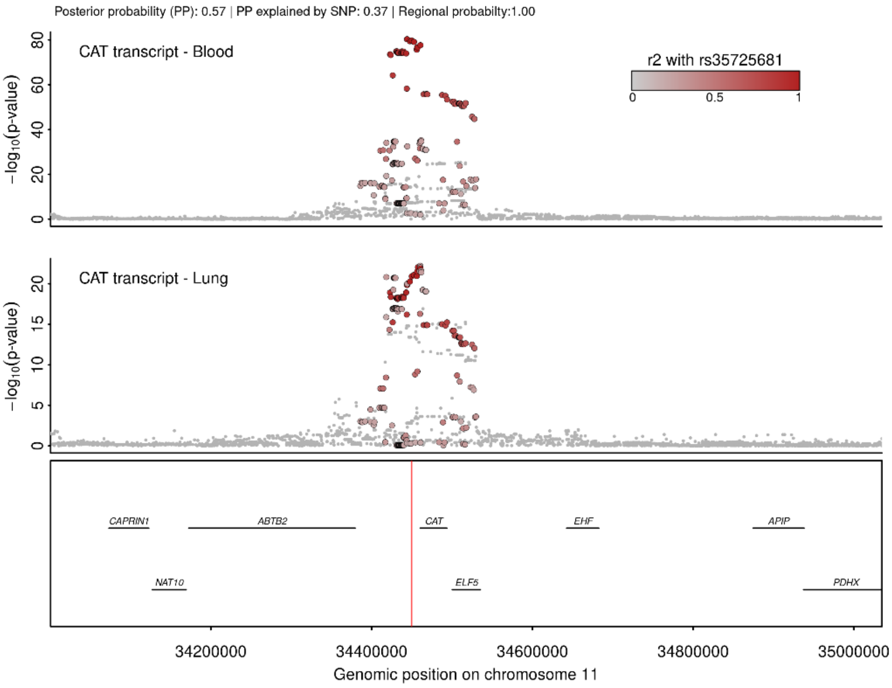


**Supplementary Figure 3 Stacked regional association plots at *CAT*.** Each panel contains regional association statistics (p-values) for the trait listed in the upper left corner along genomic coordinates. Each dot represents a single nucleotide polymorphisms and colours indicate linkage disequilibrium (LD; $r^2$) with the most likely causative variant (rs35725681) at this locus (darker colours stronger LD).

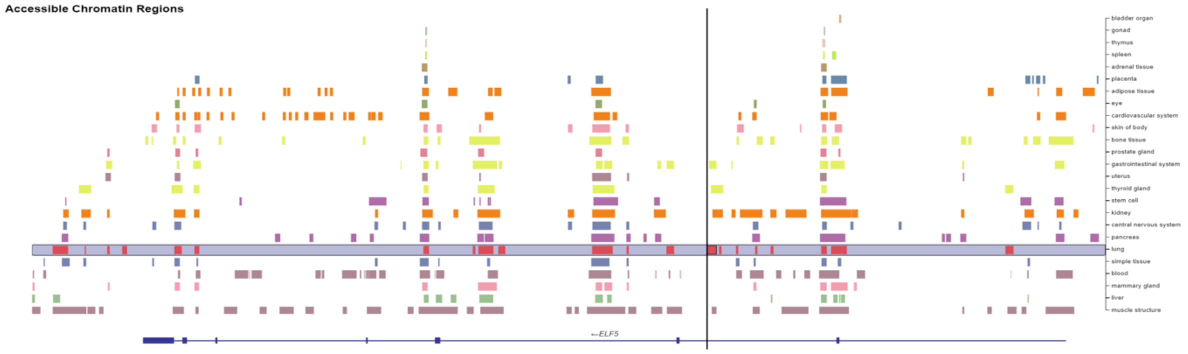


**Supplementary Figure 4 Open chromatin regions at ELF5 across various tissues.** Each colour bar indicates open chromatin regions identified using ATAC-seq experiments. The relevant lung-specific region is highlighted. Data was obtained from https://t2d.hugeamp.org/variant.html?variant=rs766826 and open chromatin regions in lung are based on experiments in alveolar type 2 cells (https://cmdga.org/annotations/DSR063NOE/). The position of rs766826 is highlighted by a black bar.

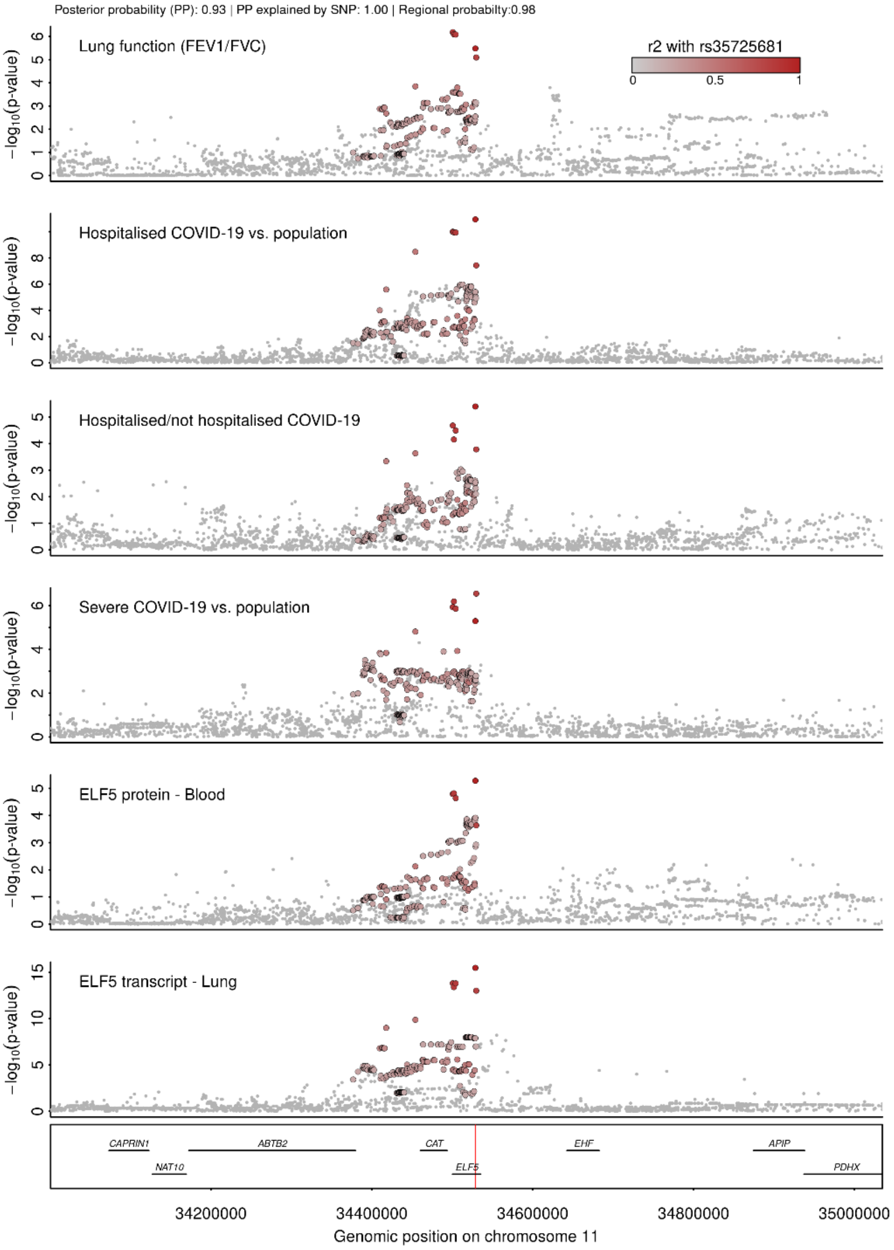


**Supplementary Figure 5 Stacked regional association plots at *ELF5*.** Each panel contains regional association statistics (p-values) for the trait listed in the upper left corner along genomic coordinates. Each dot represents a single nucleotide polymorphisms and colours indicate linkage disequilibrium (LD; r²) with the most likely causative variant (rs766826) at this locus (darker colours stronger LD). This figure is similar to Figure 2 in the main text, but now also including summary statistics for the FEV1/FCV ratio as a measure of lung function taken from Shrine et al.[1].

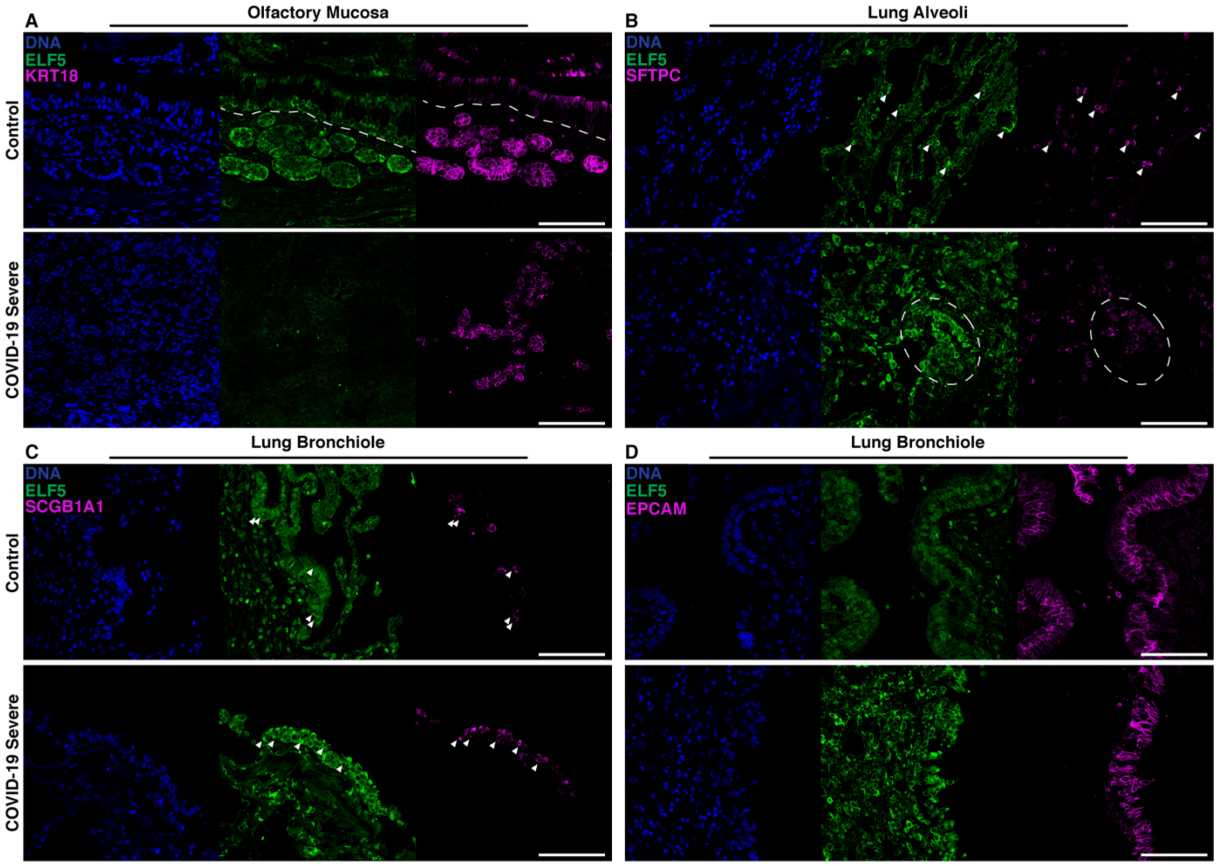


**Supplementary Figure 6 ELF5 expression by epithelial cells of the olfactory mucosa and lung.** Immunofluorescent staining of ELF5 in separate channels in control and COVID-19 patients in the **A** olfactory mucosa, **B** lung alveoli, and **C** lung bronchiole. **A** Dashed lines separate the olfactory epithelium and the lamina propria. **B** Arrowheads highlight AT2 cells expressing ELF5; dashed outline highlights clusters of AT2 cells expressing ELF5. **C** left: Arrowheads highlight secretory cells expressing ELF5; right: arrowheads highlight airway epithelial cells expressing ELF5. Marker genes for sustentacular and Bowman gland cells (**A**, KRT18), alveolar type II cells (**B**, SFTPC), secretory cells (**C**, SCGB1A1), and epithelial cells (**C**, EPCAM) are shown in purple. Validation staining for each tissue: control (n = 2); COVID-19 (n = 2). Scale bar = 100µm.

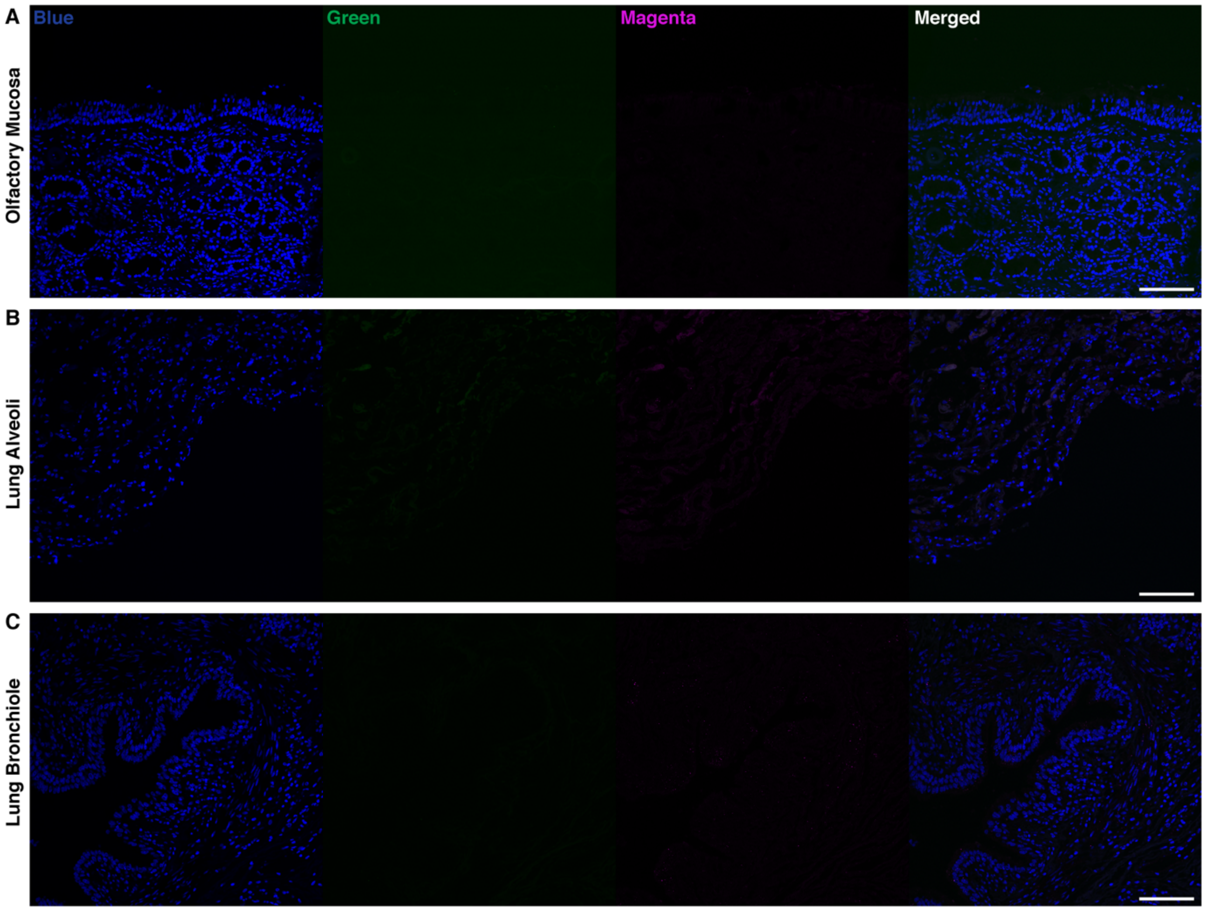


**Supplementary Figure 7. No primary antibody controls of the olfactory mucosa and lung.** Representative images of negative controls for the **A** olfactory mucosa, **B** lung alveoli, and **C** lung bronchiole under high laser power. Scale bar = 100µm.

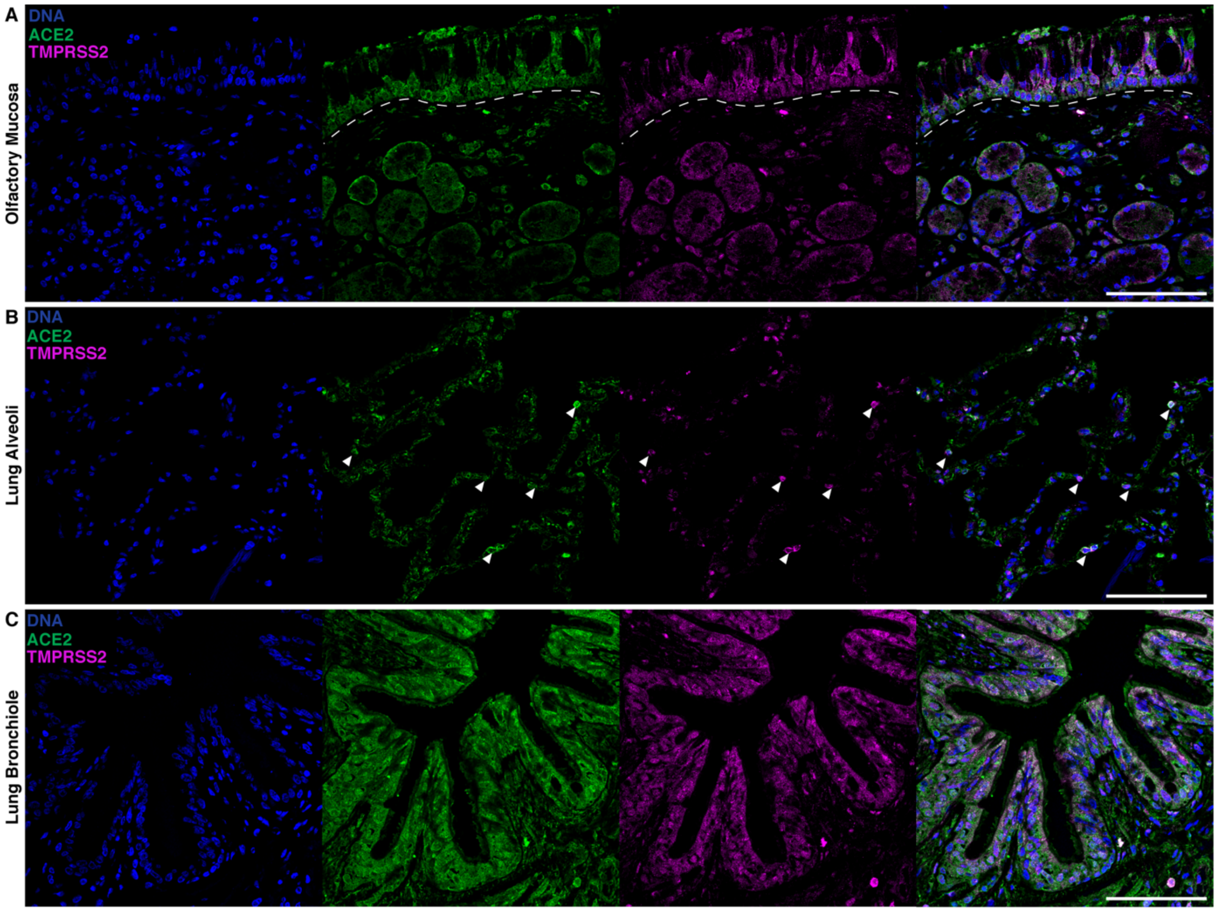


**Supplementary Figure 8. ACE2 and TMPRSS2 are expressed by different epithelial cells of the olfactory mucosa and the lung. A** Sustentacular cells, horizontal basal cells, and Bowman gland cells of the olfactory mucosa co-express ACE2 and TMPRSS2. **B** Arrows highlight punctuated expression of TMPRSS2 reflecting the distribution of AT2 cells together with ACE2. **C** Airway epithelial cells of the bronchioles expressing ACE2 and TMPRSS2. Validation staining for each tissue: control (n = 2). Scale bar = 100µm.

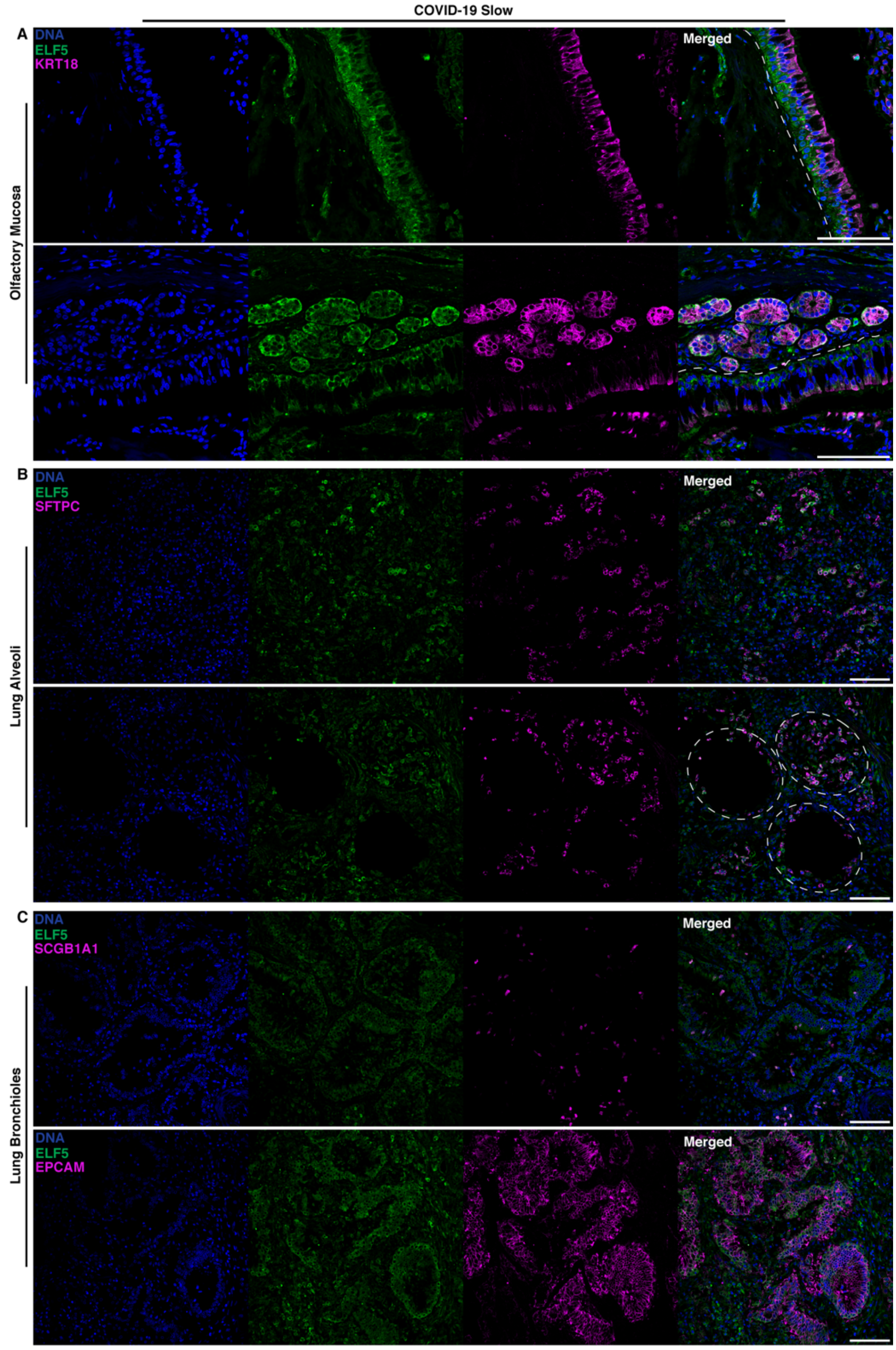


**Supplementary Figure 9. ELF5 expression by epithelial cells of the olfactory mucosa and lung in COVID-19 cases with longer disease duration.** Immunofluorescent staining of ELF5 in separate channels of COVID-19 patients that died after hospitalization after 14 days. ELF5 expression in the **A** olfactory mucosa, **B** lung alveoli, and **C** lung bronchiole. **A** Dashed lines separate the olfactory epithelium and the lamina propria. **B** dashed outline highlights clusters of AT2 cells expressing ELF5. Marker genes for sustentacular and Bowman gland cells (**A**, KRT18), alveolar type II cells (**B**, SFTPC), secretory cells (**C**, SCGB1A1), and epithelial cells (**C**, EPCAM) are shown in purple. Validation staining for each tissue: control (n = 2); COVID-19 (n = 2). Scale bar = 100µm.

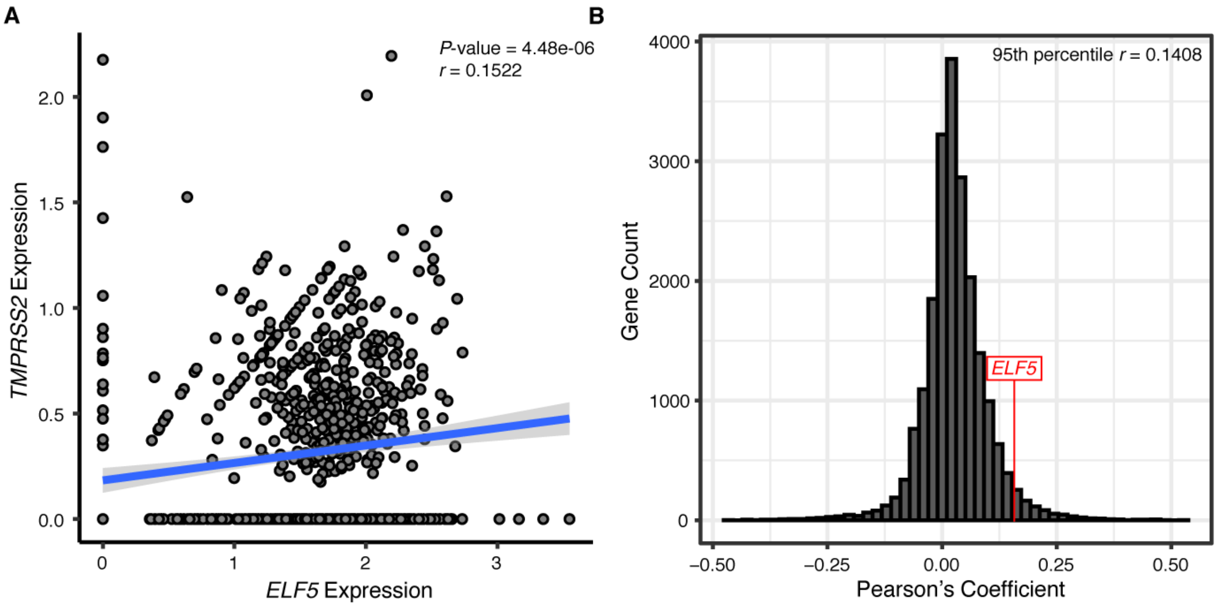


**Supplementary Figure 10. Co-expression of ELF5 and TMPRSS2.** A Scatterplot opposing normalised expression levels of ELF5 and TMPRSS2 in sustentacular cells of the olfactory mucosa. The blue line indicates a linear regression fit and correlation coefficient and p-value are given in the legend. B Distribution of correlation coefficients of pairwise gene expression across all genes detected in sustentacular cells. The red line indicates were the ELF5 – TMPRSS2 correlations is placed.

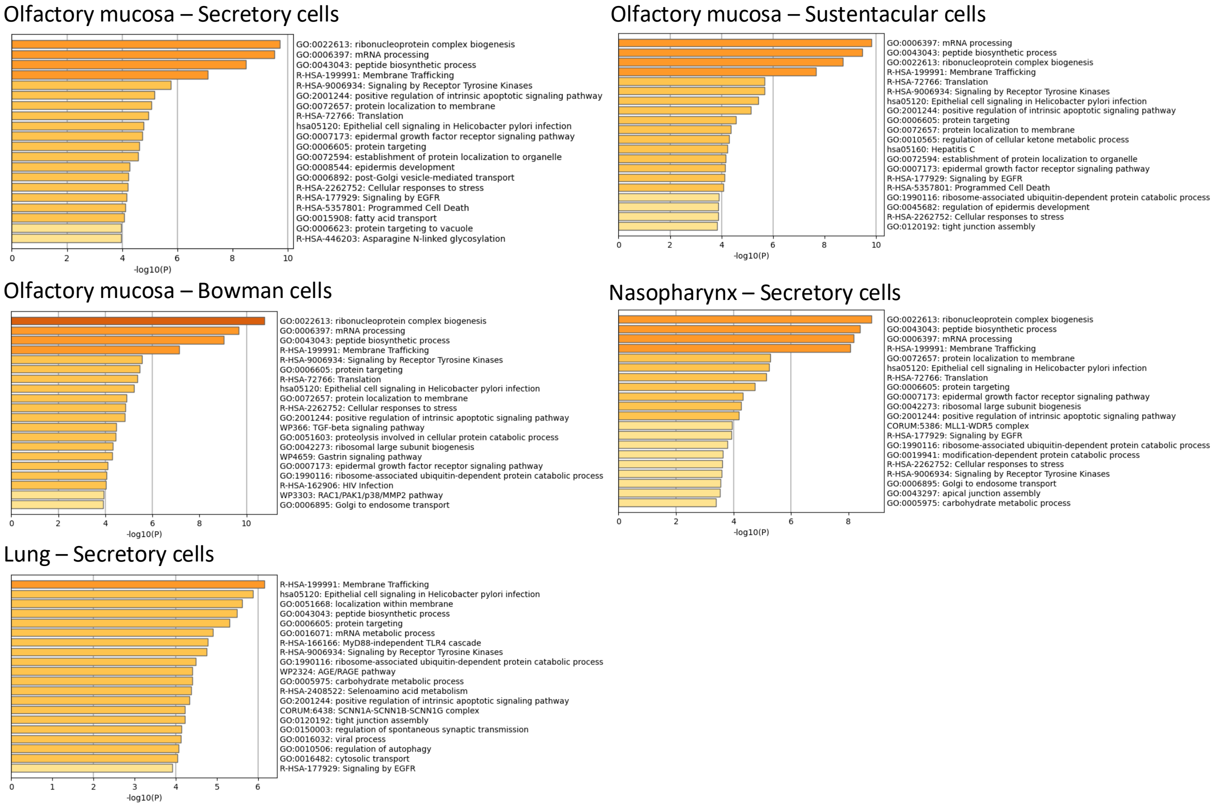


**Supplementary Figure 11.** Results from cell-type specific pathway enrichment analysis for predicted ELF5 targets.

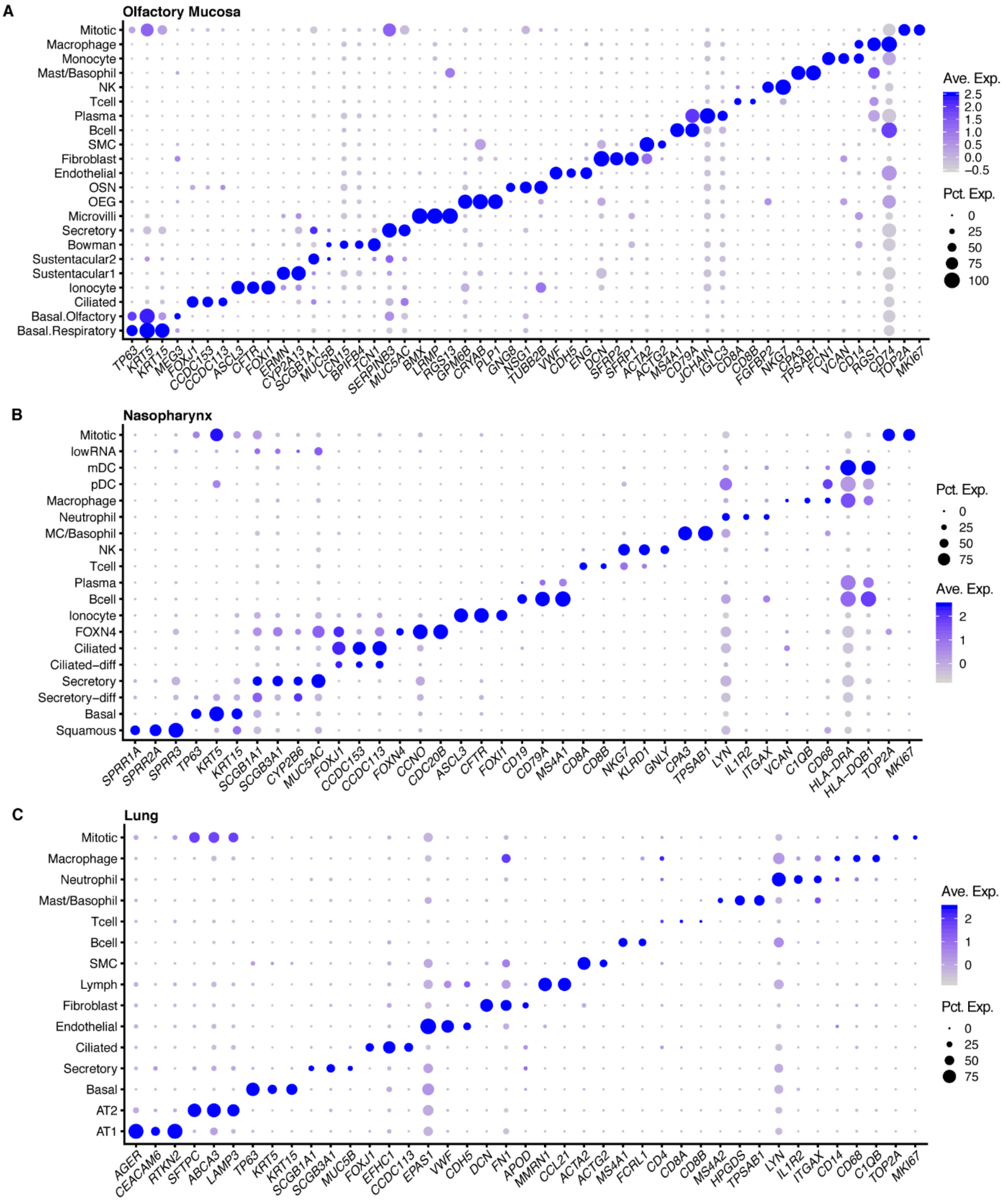


**Supplemental Figure 12.** Cell-specific expression of marker genes used for single cell annotation across the three tissue data sets used. Higher expression levels are indicated by darker shades and the size of the circle reflects the percentage of cells expressing the corresponding marker gene.
